# Performance evaluation of RespiCast ensemble forecasts for primary care syndromic indicators of viral respiratory disease in Europe

**DOI:** 10.1101/2025.10.30.25339155

**Authors:** Nicolò Gozzi, Corrado Gioannini, Paolo Milano, Ivan Vismara, Luca Rossi, Marco Quaggiotto, Valeria Marras, Stefania Fiandrino, Mattia Mazzoli, Daniela Paolotti, Alessandro Vespignani, Francesco Celino, Lorenzo Zino, Alessandro Rizzo, Sasikiran Kandula, Birgitte Freiesleben de Blasio, Maikel Bosschaert, Steven Abrams, Niel Hens, Atte Aalto, Daniele Proverbio, Giulia Giordano, Jorge Goncalves, Katharine Sherratt, Rhys Earl, Kelsey E. Shaw, T. Alex Perkins, Yuhan Li, Nicola Perra, Fuminari Miura, Don Klinkenberg, Rok Grah, Helen Johnson, Ajibola Omokanye, Leah J. Martin, Rene Niehus, Jose Canevari, Eva Bons

## Abstract

In 2023 the European Centre for Disease Prevention and Control (ECDC) launched RespiCast, the first European Respiratory Diseases Forecasting Hub, to provide probabilistic forecasts for influenza-like illness (ILI) and acute respiratory infection (ARI) incidence across 26 European countries. During the 2023/24 and 2024/25 winter seasons, RespiCast collected one- to four-week-ahead forecasts from multiple models contributed by different international teams and combined them into an ensemble. Our analysis shows that, when evaluated using the weighted interval score (WIS) and the absolute error (AE), the ensemble consistently outperformed the baseline model (defined as a persistence model that projects the last observed value forward) as well as individual models across most countries and forecasting rounds for both ILI and ARI incidence in the two seasons. Analysis of ensemble coverage (defined as the proportion of times observed values fall within the specified prediction intervals) indicated that forecast prediction intervals were reliable, although a general overconfidence trend (i.e., prediction intervals that are too narrow) was observed, particularly in specific countries. The relative performance of the ensemble declined in certain weeks, likely due to reduced participation from modelling teams, epidemic dynamics, higher data noise, and reporting delays. Forecast scores varied across countries, with some exhibiting consistently higher relative errors than others. Overall, the findings highlight the strengths of ensemble approaches in improving the accuracy and reliability of epidemiological forecasts while identifying areas for improvement, such as managing overconfidence and addressing variability in performance across countries and over time.

## 1. Introduction

Infectious disease forecasting can play a critical role in informing health authorities about the dynamics of epidemic outbreaks, guiding the timely implementation of public health measures and effective communication strategies, as well as optimizing resource allocation [1], [2]. In this context, collaborative forecasting hubs have emerged as an effective strategy to approach the challenges linked to infectious disease forecasting [3], [4]. In particular, they address the characterization of uncertainty in disease modelling by integrating different approaches which capture different aspects of epidemic dynamics. Since no single model is likely to account for all the biological, behavioural, and environmental factors influencing an outbreak, combining multiple model outputs allows for a more comprehensive representation of potential epidemic trajectories.

The forecasting hub approach brings together diverse modelling teams with the goal of creating ensemble forecasts (i.e., combined predictions from multiple models) that, in past efforts, have consistently outperformed the predictions of individual models across multiple scoring criteria [5], [6], [7]. Besides their operational focus, these hubs also serve as collaborative research platforms to test novel forecasting approaches, develop new model evaluation and ensembling techniques, and promote continuous scientific advancement through shared insights.

During the COVID-19 pandemic, the European Centre for Disease Prevention and Control (ECDC) introduced the European COVID-19 Forecasting Hub that supported the EU’s management of the health crisis [5]. Since 2021, this hub has provided real-time forecasts of cases, deaths, and hospitalizations related to COVID-19, with the aim of supporting public-health decision making. As we moved out of the acute phases of the COVID-19 pandemic, Europe shifted toward an integrated surveillance system for respiratory pathogens, now monitoring multiple viruses within a unified framework. A concrete example of this tendency is the European Respiratory Virus Surveillance Summary (ERVISS), a joint effort between ECDC and the WHO Regional Office for Europe that provides weekly integrated summaries of the epidemiological and virological situation for influenza, respiratory syncytial virus (RSV), and SARS-CoV-2 in Europe, based on national surveillance data submitted to The European Surveillance System (TESSy) [8]. This approach to integrated respiratory pathogen surveillance calls for a similarly integrated strategy in forecasting; namely, the establishment of a unified European forecasting hub for viral respiratory diseases, capable of producing forecasts across multiple pathogens.

In this study, we present the structure and results of RespiCast, a new European forecasting hub for respiratory diseases, launched by ECDC during the 2023/24 winter season. As forecasting targets, in its first seasons RespiCast focused on primary care *epidemiological* indicators—specifically influenza-like illness (ILI) and acute respiratory infection (ARI) incidence—to provide a broad representation of viral respiratory activity in the community. RespiCast conducted 20 and 28 forecasting rounds during the 2023/24 and 2024/25 seasons, respectively, aggregating predictions from multiple modelling teams for ILI and ARI incidence across 26 European countries. We evaluate the performance of the ensemble forecasts generated by RespiCast in its first two seasons by comparing them to individual models and to a naive baseline using the weighted interval score (WIS), the absolute error (AE), and prediction coverage, and we examine how forecast accuracy varies over time and across different geographical contexts.

## 2. Methods

### 2.1 Description of the RespiCast forecasting hub

Starting from the 2023/24 respiratory virus season, modelling teams worldwide were invited to participate in the RespiCast forecasting challenge. The challenge lasted for 20 rounds (i.e., forecast submission cycles) from 2023/12/20 to 2024/05/01 in the 2023/24 season and for 28 rounds from 2024/10/23 to 2025/04/30 in the 2024/25 season (more details on contributing teams are provided in the Supplementary Information). Data were typically updated on Fridays, covering the previous week (i.e., up to the preceding Sunday), and teams were required to submit their forecasts by the following Wednesday. These forecasts were then combined into an ensemble and published on the RespiCast website [9] every Thursday (see Figure 1 for the representation of the workflow of a RespiCast forecasting round). The forecasting horizons ranged from 1 to 4 weeks ahead. Given the timing of the data updates, the forecasts published on Thursdays covered the preceding week (for which public data had not yet been consolidated), the current week, and the two subsequent weeks. More formally, let *r* denote a forecasting round and *t*_*r*_ the reference time for that round, corresponding to the most recent epidemiological week for which data was available at the time of submission. For each round *r*, participating teams submitted probabilistic forecasts for multiple horizons *h* ∈ {1, 2, 3, 4} weeks ahead. The target time for a forecast at horizon *h* is then *t*_*r*_ + *h*, and the corresponding observed outcome (e.g., reported ILI incidence) is denoted by 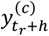 for country *c*. All forecast distributions 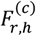 are constructed to predict these future outcomes. Only probabilistic forecasts in the form of quantiles were accepted, and teams were allowed to submit forecasts for one or multiple countries. Full details on the submission format and requirements are provided on the GitHub RespiCast repository [10], [11].

**Figure 1.**
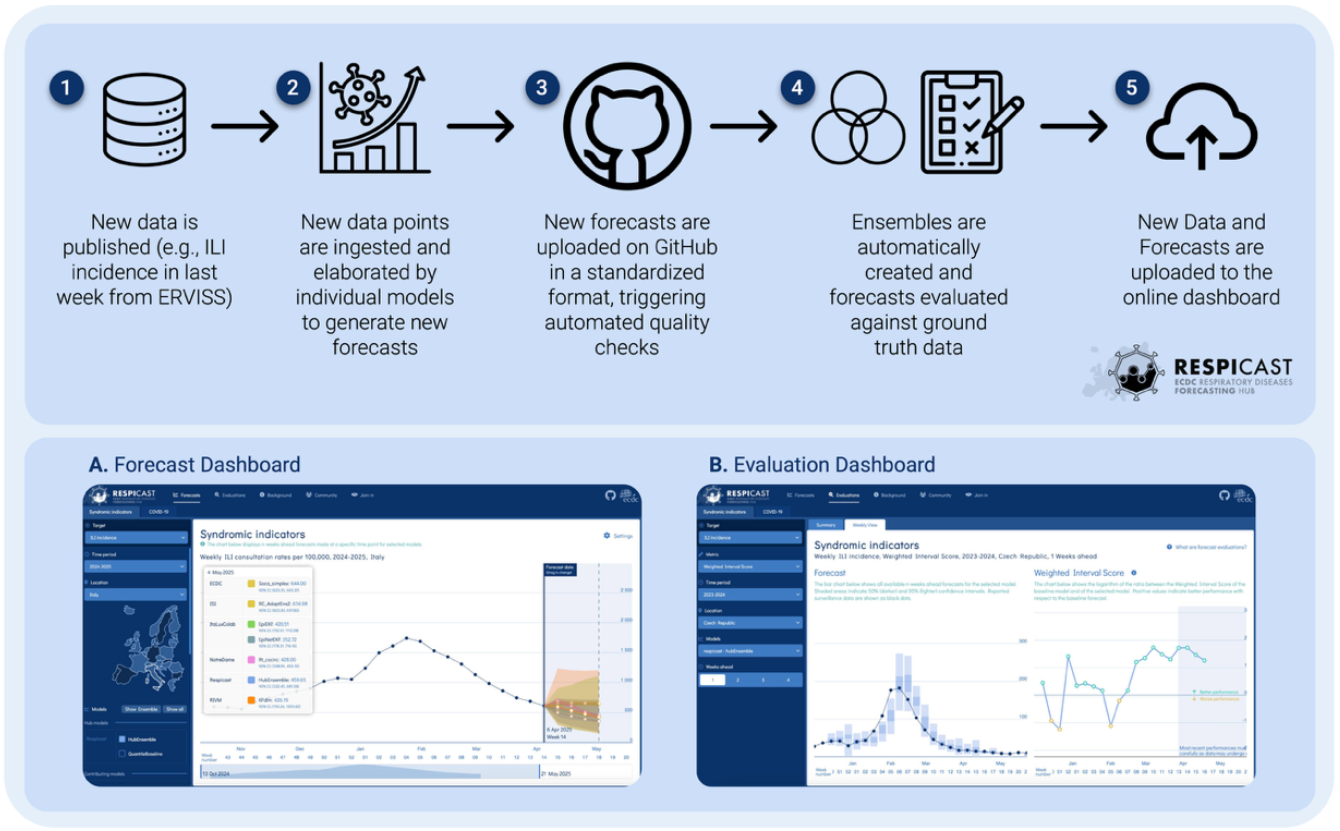
RespiCast forecasting round workflow (top row) and overview of the forecasting platform components (bottom row).

### 2.2 Forecasting targets

Teams were asked to submit probabilistic forecasts on both ILI and ARI incidence, expressed as reported cases per 100,000 population units in all countries except for Cyprus, Luxembourg, Malta, and Finland where rates are expressed per 100,000 consultations. Both ILI and ARI are syndromic indicators typically estimated through sentinel primary care surveillance. Specific case definitions can be found in [12].

Historical data since the 2014/15 season was made available to the modelling teams, and participants were also allowed to use additional data sources of their choice to inform their models (e.g., serological, social contact data). ILI incidence was available for 22 European countries, while ARI incidence was available for 16 countries. Data for all countries was sourced from the European Respiratory Virus Surveillance Summary (ERVISS) [8]. We note that RespiCast also accepts forecasts for ILI and ARI incidence for the countries of the United Kingdom and Switzerland. Data for these countries are sourced from FluID [13] and were added during the course of the 2023/24 season. During 2024/25, a disruption in FluID data imports resulted in missing observations for most rounds for these countries. For this reason, we excluded them from the performance analysis.

Multiple pathogens contribute to ILI and ARI incidence rates. However, they differ in their sensitivity and specificity for representing various respiratory pathogens, which supports the use of both as forecasting targets to better track the progression of respiratory diseases in Europe. The ARI case definition, for instance, does not include high temperature, making it generally more sensitive to a wider range of pathogens like SARS-CoV-2 and RSV. However, this more inclusive case definition comes at the cost of lower specificity compared to ILI, particularly for influenza [12].

### 2.3 Ensemble and baseline forecasting model

RespiCast combines multi-model projections into an ensemble forecast considering a median-across-quantiles approach [14]. For each country *c*, round *r*, and horizon *h*, let 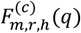 denote the cumulative distribution function of forecast submitted by model *m*. It follows that 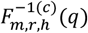 denote the predictive quantile of level *q*. The ensemble forecast is defined as the quantile-wise median across models:

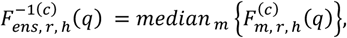

where 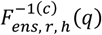 is the ensemble’s predictive quantile at level *q*.

The baseline is a naive, largely uninformed model which serves as a neutral benchmark for model evaluation. Across all time horizons, the baseline model predicts the last data point within the calibration period as its median, while the prediction intervals are based on past data increments. The baseline forecast will thus always be a flat line with symmetric prediction intervals reflecting country-specific historical variation in the data. More precisely, for each country *c*, the baseline forecast for round *r* and horizon *h* is constructed by extrapolating forward from the most recently observed data point 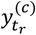, where *t*_*r*_ is the reference time of the forecast round (i.e., the time with the most recent available data point). The baseline assumes future changes follow the empirical distribution of past 1-week differences. We define the set of past weekly increments in the target quantity (e.g., ILI incidence) as: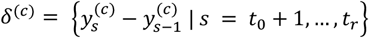, where *t*_0_ is the start of the baseline calibration window (week 42 of 2023). To ensure that the median baseline forecast aligns with the last calibration point, we symmetrize *δ* by considering 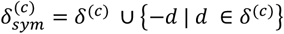. From this symmetric set, we sample with replacement *H* increments (*d*_1_, …, *d*_*H*_) to construct predictive trajectories. The value of each trajectory at horizon *h* is computed as 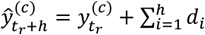, where *h* ∈ {1, ⋯, *H*} and *t*_*r*_ is the reference time of the forecast round. Negative values possibly resulting from this procedure are set automatically to zero. We repeat this process to generate *N* = 10,000 synthetic trajectories, from which we compute forecast quantiles of the baseline 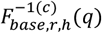. Analogous baseline modelling approaches have been employed in other epidemiological forecasting challenges [5], [6], [15].

### 2.4 Forecast evaluation

Forecasts are evaluated against reported surveillance data as of October 24, 2025. We assess predictive performance using the principle of maximizing the sharpness of predictive distributions while maintaining proper calibration. This means that prediction intervals should be as narrow as possible and closely aligned with the real data, while ensuring that forecasted probabilities accurately reflect observed frequencies. The overall predictive performance is quantified using both the WIS and the AE of the median, while we assess calibration by means of the prediction coverage. Relative performance is expressed by means of the relative WIS and relative AE. In the next subsections, we briefly introduce these concepts.

#### Weighted interval score (WIS)

The WIS is a scoring rule used to evaluate the accuracy of probabilistic forecasts [16]. It assesses the performance of quantile-based forecasts by penalizing both the distance between predicted and actual values, as well as under- or over-prediction. For a (1 − *α*) × 100% prediction interval of a forecast *F*, the interval score (IS) is defined as:

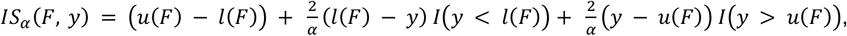

where *y* is the observed outcome, *u*(*F*) is the upper limit of the prediction interval of forecast *F, l*(*F*) is the lower limit thereof, and *I*(·) is an indicator function that equals one when the specified condition is true, and zero otherwise. It follows that the first term measures the width of the prediction interval, while the second and third terms are penalties for, respectively, over- and under-prediction of the forecast weighted by the inverse of level *α*. The WIS is an extension of this metric to multiple intervals. More specifically, for *K* (1 − *α*_*k*_) × 100% prediction intervals (*k* = 1, …, *K*) of forecast *F*, the WIS is defined as:

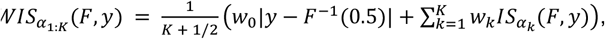

where *F*^−1^(0.5) is the projected median and *w*_0_, *w*_*k*_ are non-negative weights that we set, following a standard choice, to 1/2 and *α*_*k*_/2, respectively. Here, we consider *K* = 11 prediction intervals (*α*_*k*_ = 0.02, 0.05, 0.10, 0.20, 0.30, 0.40, 0.50, 0.60, 0.70, 0.80, 0.90). The WIS is computed for each forecast submission 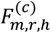 and the corresponding target outcome 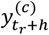, where *r* is the forecasting round, *h* is the forecast horizon, and *c* denotes the country.

#### Absolute error (AE)

The AE is a commonly used metric to evaluate the accuracy of point forecasts. It measures the absolute difference between the predicted and the observed value, indicating how far off a prediction is from the reported data. Given a forecast distribution *F* and observed value *y*, the AE is defined as:

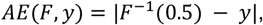

where *F*^−1^(0.5) denotes the median of the forecast distribution. Note that this metric measures the absolute distance between the median projection and the observed value in contrast with the WIS that also accounts for forecast uncertainty. In our framework, the AE is computed for each forecast submission 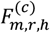and the corresponding target outcome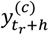.

#### Prediction coverage

Prediction coverage is a metric used to evaluate calibration of probabilistic forecasts, specifically by measuring the proportion of times the observed values fall within the prediction intervals. For (1 − *α*) × 100% intervals, the prediction coverage over *N* forecast-observation pairs (*F*_*i*_, *y*_*i*_) is defined as:

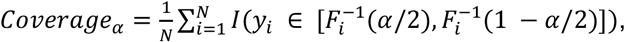

where *y*_*i*_ are the observed values, 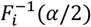 and 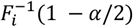 are the lower and upper bounds of the (1 − *α*) × 100% prediction interval for the i-th forecast, and *I*(·) equals one when the observed value falls within the prediction interval and zero otherwise.

A well-calibrated model will have prediction coverage values closely matching the nominal coverage (e.g., 90% of values fall in the 90% prediction interval), while deviating values indicate over- or under-confidence. A model is overconfident if the prediction coverage is below the nominal coverage: in this case, the prediction interval is too narrow (e.g. only 80% of values fall in the 90% prediction interval). The reverse is true for an underconfident model: the prediction coverage is above the nominal coverage meaning the prediction interval is *too* wide.

#### Relative performance

To evaluate the performance of a model *m* relative to the baseline, we define the relative WIS as the base-2 logarithm of the ratio between the WIS of the baseline model and that of model *m*. For a given forecasting round *r*, horizon *h*, and country *c*, it is defined as:

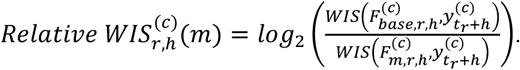

Positive values indicate that the model performs better than the baseline, while negative values indicate worse performance relative to the baseline. An analogous definition can be applied to the AE of the median. Unless stated differently, relative performance across different horizons within the same forecasting round is averaged.

### 2.5 RespiCast forecasting platform

The RespiCast infrastructure consists of two primary components: a set of GitHub repositories with automated actions and a dedicated web platform focused on forecast visualization and evaluation [9]. For each set of disease indicators, a corresponding repository is created and configured on GitHub. These repositories store ground truth data for model calibration and evaluation and support the submission and validation of forecasts from contributing teams. The repository structure adheres to the recommendations of the *hubverse* standard, a collection of open-source software and tools that support collaborative modelling challenges [17]. The RespiCast web platform features two interactive applications for exploring forecasts and evaluating model performance. The forecast app allows users to select targets, seasons, locations, models and visualize reported data along with forecasts. The evaluations app has a similar structure, enabling users to choose targets, metrics, and seasons and to explore forecasts performance. Figure 1 provides a snapshot of the two dashboards.

## 3. Results

### 3.1 Participation and engagement

During the 2023/24 winter season, RespiCast collected forecasts for ILI and ARI incidence over 20 rounds in 26 European countries. As shown in Table 1, by the end of the season, 14 different forecasting models from 6 contributing teams were submitted to forecast ILI incidence, while results from 10 models from 3 teams were submitted for ARI incidence. On average, the outputs of 11 models were submitted per round for ILI incidence, and from 7 models for ARI incidence. The forecasts covered 22 European countries for ILI and 14 countries for ARI incidence. The 2024/25 season saw increased participation. The forecasting season lasted 28 rounds, during which 21 models from 9 teams were submitted for ILI incidence, and 14 models from 6 teams for ARI incidence. Participation at each round also increased, with 15 ILI models and 8 ARI models submitted on average each week. For a more detailed overview of the number of models over time, see Figure S1 in the Supplementary Information.

**Table 1:**
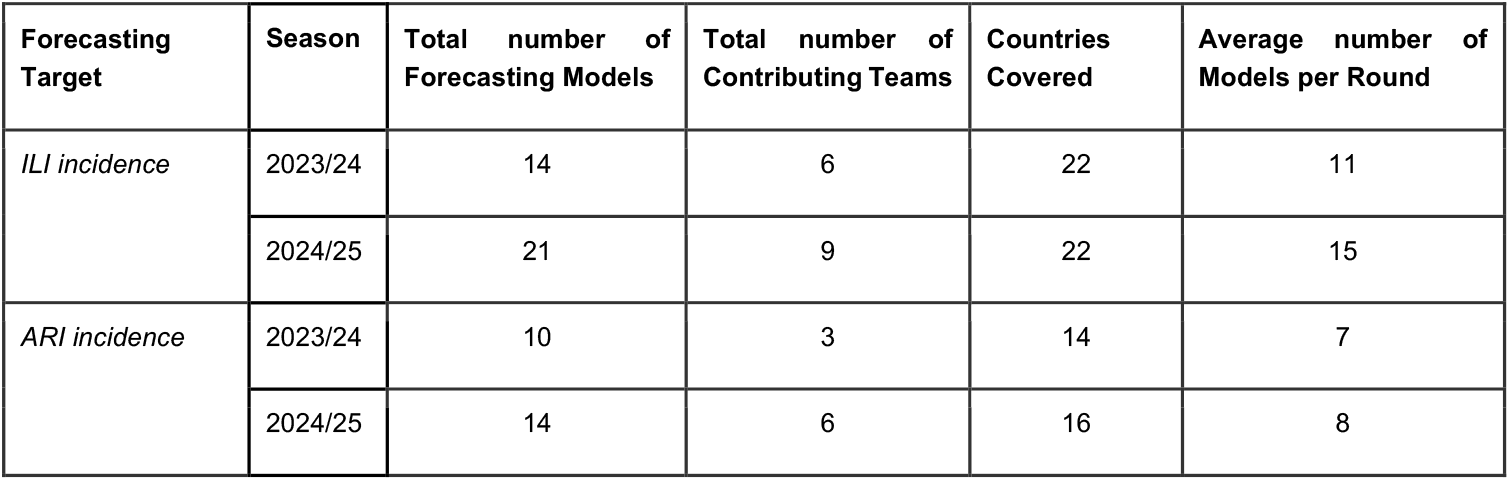
Summary of contributions to RespiCast for each forecasting target during 2023/24 and 2024/25 forecasting seasons.

| Forecasting Target | Season | Total number of Forecasting Models | Total number of Contributing Teams | Countries Covered | Average number of Models per Round |
| --- | --- | --- | --- | --- | --- |
| ILI incidence | 2023/24 | 14 | 6 | 22 | 11 |
|  | 2024/25 | 21 | 9 | 22 | 15 |
| ARI incidence | 2023/24 | 10 | 3 | 14 | 7 |
|  | 2024/25 | 14 | 6 | 16 | 8 |

In the following analysis, we focus on rounds and countries where at least three distinct models (excluding the ensemble and baseline) were submitted. This ensures that ensembles were generated from a minimum of three models in each round considered. Additionally, forecasts for Latvia between 2024/04/21 and 2025/02/02 were excluded due to anomalies in the reported data in this period, where incidences were consistently reported as 100,000 cases per 100,000 individuals. This leads to the inclusion of 22 countries for ILI incidence in both seasons, and 14 and 16 for ARI incidence in 2023/24 and 2024/25 seasons, respectively. It is important to note that not all countries reported surveillance data with the same frequency and that some did not report data during certain weeks. No data was reported in week 52 due to the end-of-year holidays in both seasons.

In the Supplementary Information, we provide a description of submitted models for each target and season. The sample includes a diverse range of approaches, including statistical methods based on autoregressive models such as ARIMA, fully mechanistic models incorporating country-specific demographics and contact patterns, metapopulation models that account for individual mobility across and within countries, and semi-mechanistic approaches based on effective reproductive number estimation, generalized logistic growth, and data integration techniques such as the extended Kalman filter. Additionally, some models employ weighted ensembles of individual approaches which are not among the models submitted independently by other participants.

### 3.2 Forecasting performance during 2023/24 and 2024/25 winter seasons

Figure 2 illustrates the performance of ensemble forecasts for ILI incidence using the relative WIS (i.e., relative to the baseline model). Figure 2A presents the distribution of relative WIS values, aggregating results across all countries and forecasting rounds, separately for the ensemble and all other models in both seasons. In the 2023/24 season, pooling together scores from individual models we obtain a median relative WIS of 0.08, indicating marginal improvements over the baseline. In contrast, the ensemble’s median relative WIS is 0.44, demonstrating superior predictive performance as compared to the baseline and the pool of individual models. The ensemble exhibits also greater stability and consistency across forecasting rounds and countries. In fact, its distribution of relative WIS values is narrower, with an interquartile range (IQR) of 1.25 compared to 1.51 for the pool of individual models. Similar patterns are observed in the 2024/25 season, with the ensemble’s median relative WIS at 0.40 compared to 0.06 for individual models, and an IQR of 0.91 versus 1.28.

**Figure 2.**
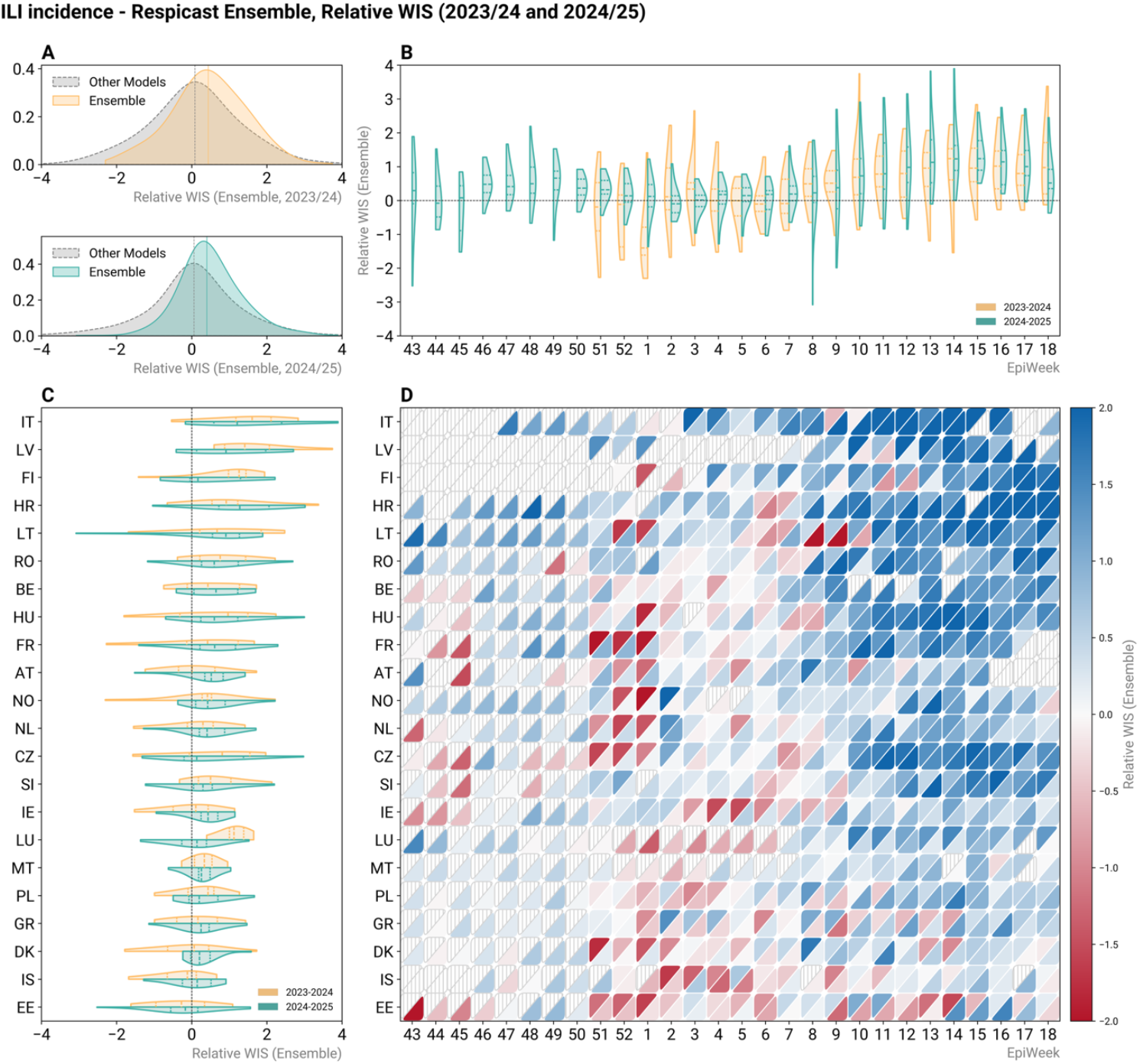
Relative Weighted Interval Score (WIS) for Influenza-Like Illness (ILI) incidence predictions from the RespiCast Ensemble during 2023/24 and 2024/25 winter seasons. **(A)** Distribution of relative WIS across all countries and weeks, comparing the ensemble model to all other models combined in 2023/24 (top) and 2024/25 (bottom) winter seasons. **(B)** Distribution of relative WIS of the ensemble across all countries for each epidemiological week separately (2023/24 in yellow, left half; 2024/25 in green, right half of the split violin). **(C)** Distribution of relative WIS of the ensemble across all epidemiological weeks for each country separately (2023/24 in yellow, top half; 2024/25 in green, bottom half of the split violin). **(D)** Heatmap of relative WIS values of the ensemble by country and epidemiological week, with blue indicating better performance (relative WIS greater than 0) and red indicating worse performance (relative WIS lower than 0) compared to the baseline. The top-left half of each cell shows data for the 2023/24 season, while the bottom-right half shows data for 2024/25. Cells with grey stripes indicate missing data. The violin plots in Panels B and C are truncated at the observed minimum and maximum values, with dashed lines inside representing the 1^st^ quartile, median, and 3^rd^ quartile of the distribution. In all panels, relative WIS values are averaged over the four-week forecasting horizon.

Ensemble performance appears to be heterogeneous in time and space. Figure 2B shows the temporal evolution of the relative WIS across epidemiological weeks in both seasons. Here, the distribution of the relative WIS for a given week consists of relative WIS scores of forecasts produced in that week, averaged over the four-week forecasting horizon and aggregated across all countries. For 2023/24, Figure 2B reveals weaker ensemble performance compared to the baseline during the early weeks of the season, corresponding to the Christmas-New Year period. Despite these early challenges, the ensemble maintains a positive median relative WIS in all but seven weeks, underscoring overall reliability and robustness over time. In 2024/25, we observe an improved performance of the ensemble over time. The median relative WIS of the ensemble is positive in 26 of the 28 forecasting weeks. As in 2023/24, in 2024/25, we also observe a decline in performance compared to the baseline in the weeks corresponding to the Christmas-New Year period. These trends may be attributed to several factors, including fewer model submissions (in the case of 2023/24 season), reduced data availability over the Christmas-New Year period, and increased heterogeneity of the epidemic phase (see Discussion Section).

Figure 2C displays the distribution of the relative WIS of the ensemble across countries (aggregating scores across all weeks), ranked by overall median performance in both seasons. In 2023/24, the ensemble generally outperforms the baseline in 20 of the 22 countries, with a negative median relative WIS observed in only two (Estonia and Iceland). In 2024/25, Estonia is the only country in which the ensemble had a negative median relative WIS. Additionally, in 2023/24, the first quartile of the distribution of the relative WIS is positive for 10 out of 22 countries, indicating that the ensemble outperformed the baseline during at least 75% of the rounds in these countries. This number increases to 14 out of 22 in 2024/25. Finally, Figure 2D provides a more granular view of the relative WIS of the ensemble by country and week. The heatmap shows the relative WIS, averaged over the four-week horizon, of ensemble forecasts produced in each week and country in both the seasons considered. The upper-left triangle reports results for the 2023/24 season, and the lower-right for the 2024/25 season. Blue indicates better performance with respect to the baseline, and red worse performance, with color intensity proportional to the relative WIS value. The figure emerging confirms distinct spatial and temporal performance patterns. Forecasts in some countries (e.g., Croatia, Finland, Italy, Latvia) consistently show positive relative WIS across nearly all weeks, while others (e.g., Belgium, France, Norway) exhibit challenges but improve over time. Conversely, in countries like Estonia and Iceland, the ensemble struggles to outperform the baseline in both seasons. Across the board, the performance improves over time in all countries, with more blue-shaded cells appearing in later weeks.

In Figure 3, we replicate the analysis for ARI incidence, revealing overall consistency with the findings for ILI. The ensemble again demonstrates performance improvements in terms of relative WIS compared to individual models (Figure 3A). However, its median relative WIS is 0.27 and 0.21 in season 2023/24 and 2024/25, respectively, lower than the values observed for ILI incidence. We note that model composition of the ensemble is different between the two targets, making the interpretation of such comparison not straightforward. Performance patterns across locations and times align with those seen for ILI incidence. In 2023/24, ensemble forecasts produced in earlier weeks exhibit lower relative WIS, with performance improving as the season progresses (Figure 3B). In 2024/25, forecasting performance also declines around the Christmas-New Year period (Figure 3B). Across countries, in 2023/24 only Finland has a negative median relative WIS, while in 2024/25 this is observed for Cyprus, Czechia, Finland, and Latvia (Figure 3C). This indicates that the ensemble outperforms, in median terms, the baseline in most locations in both seasons. Finally, Figure 3D highlights the persistent heterogeneity in patterns across locations and times, with a clear trend of improving performance over the weeks and certain countries consistently facing greater forecasting challenges, such as Cyprus and Finland.

**Figure 3.**
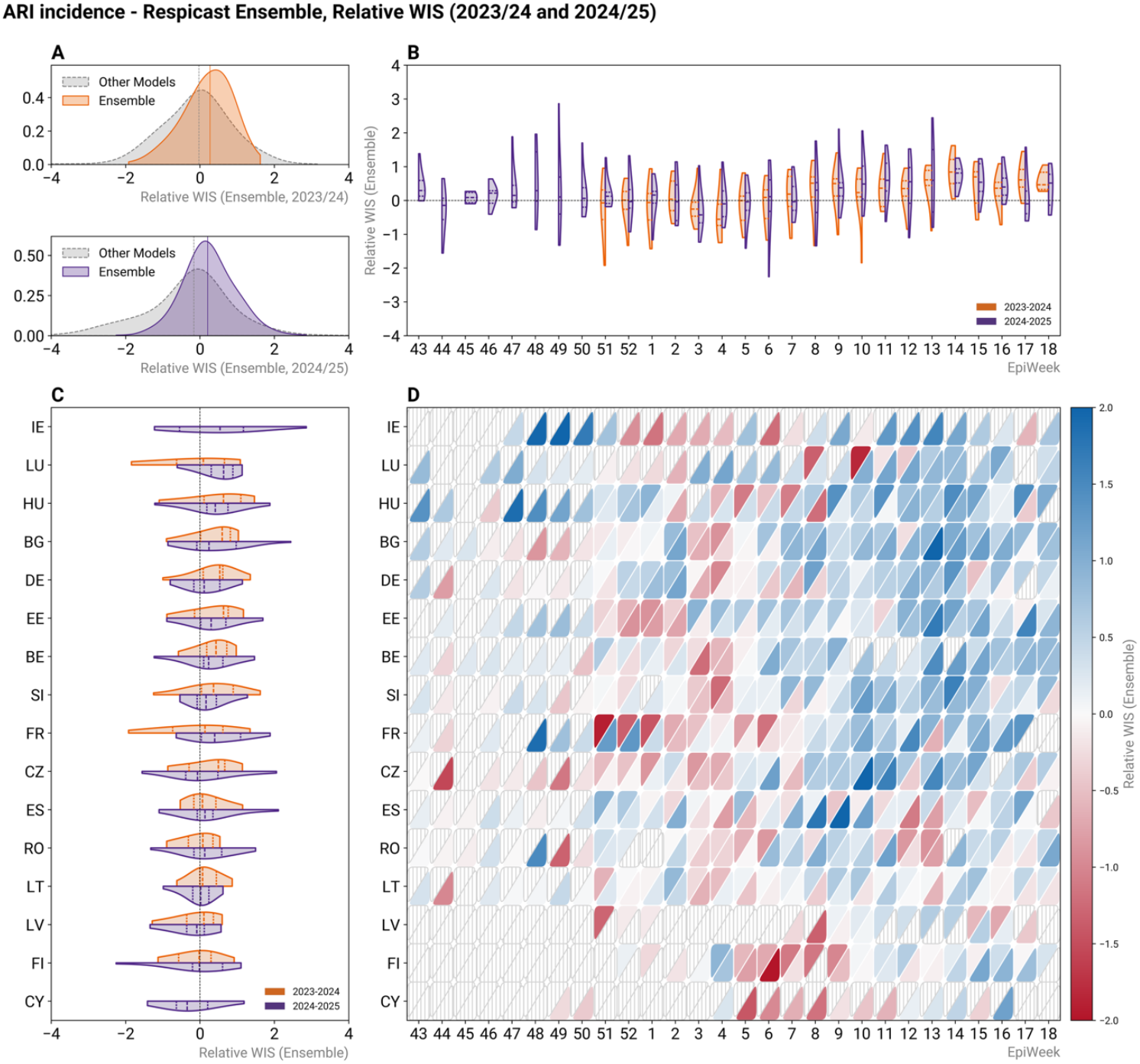
Relative Weighted Interval Score (WIS) for Acute Respiratory Infection (ARI) incidence predictions from the RespiCast Ensemble during the 2023/24 and 2024/25 winter seasons. **(A)** Distribution of relative WIS across all countries and weeks, comparing the ensemble model to all other models combined in 2023/24 (top) and 2024/25 (bottom) winter seasons. **(B)** Distribution of relative WIS of the ensemble across all countries for each epidemiological week separately (2023/24 in orange, left half; 2024/25 in purple, right half of the split violin). **(C)** Distribution of relative WIS of the ensemble across all epidemiological weeks for each country separately (2023/24 in orange, top half; 2024/25 in purple, bottom half of the split violin). **(D)** Heatmap of relative WIS values of the ensemble by country and epidemiological week, with blue indicating better performance (relative WIS greater than 0) and red indicating worse performance (relative WIS smaller than 0) compared to the baseline. The top-left half of each cell shows data for the 2023/24 season, while the bottom-right half shows data for 2024/25. Cells with grey stripes indicate missing data. The violin plots in Panels B and C are truncated at the observed minimum and maximum values, with dashed lines inside representing the 1^st^ quartile, median, and 3^rd^ quartile of the distribution. In all panels, relative WIS values are averaged over the four-week forecasting horizon.

Overall, these results highlight the ensemble’s predictive capacity while identifying specific countries and phases where the performance needs further improvement. In the Supplementary Information, we show four-week ahead ensemble forecasts for all weeks and countries, along with reported data for both targets and seasons (Figures S2-S7). We also replicate the analyses presented in Figures 2 and 3, using the relative AE instead of the relative WIS as scoring rule (Figures S12 and S13). Overall, the results remain consistent, confirming the ensemble’s improved forecasting performance compared to the baseline model. However, the magnitude of improvement is reduced when quantified using the relative AE rather than relative WIS. Additionally, in the Supplementary Information, we provide a comparison of the ensemble with individual models using both relative WIS and AE across the two forecasting targets (Figures S15-S18). The results show that the ensemble generally outperforms most individual models and has more stable and consistent performance. In particular, it most frequently ranks in the top third of the WIS or AE rank distribution across rounds and countries. In parallel, the ensemble also least frequently features among the worst-performing models in any given round or country (Figures S19 and S20).

In Figure 4 we evaluate the calibration of forecasts by comparing prediction coverage (y-axis) with nominal coverage (x-axis) across the four forecasting horizons separately (1 to 4 weeks ahead) for each forecasting target and season. Figure 4A and Figure 4B show results for ILI and ARI incidence, respectively, with the top row in each of them corresponding to the 2023/24 season and the bottom row to 2024/25. Each point represents the prediction coverage obtained by pooling together forecasts across all countries and weeks, with the dashed diagonal line indicating perfect calibration (i.e., where prediction intervals match nominal coverage). Points above the diagonal indicate underconfident forecasts, where prediction intervals are *too wide* and coverage exceeds the nominal level. Conversely, points below the diagonal suggest overconfident forecasts, where prediction intervals are *too narrow*, leading to underestimation of uncertainty and lower-than-expected coverages. The extent of deviation from the diagonal reflects the magnitude of miscalibration.

**Figure 4.**
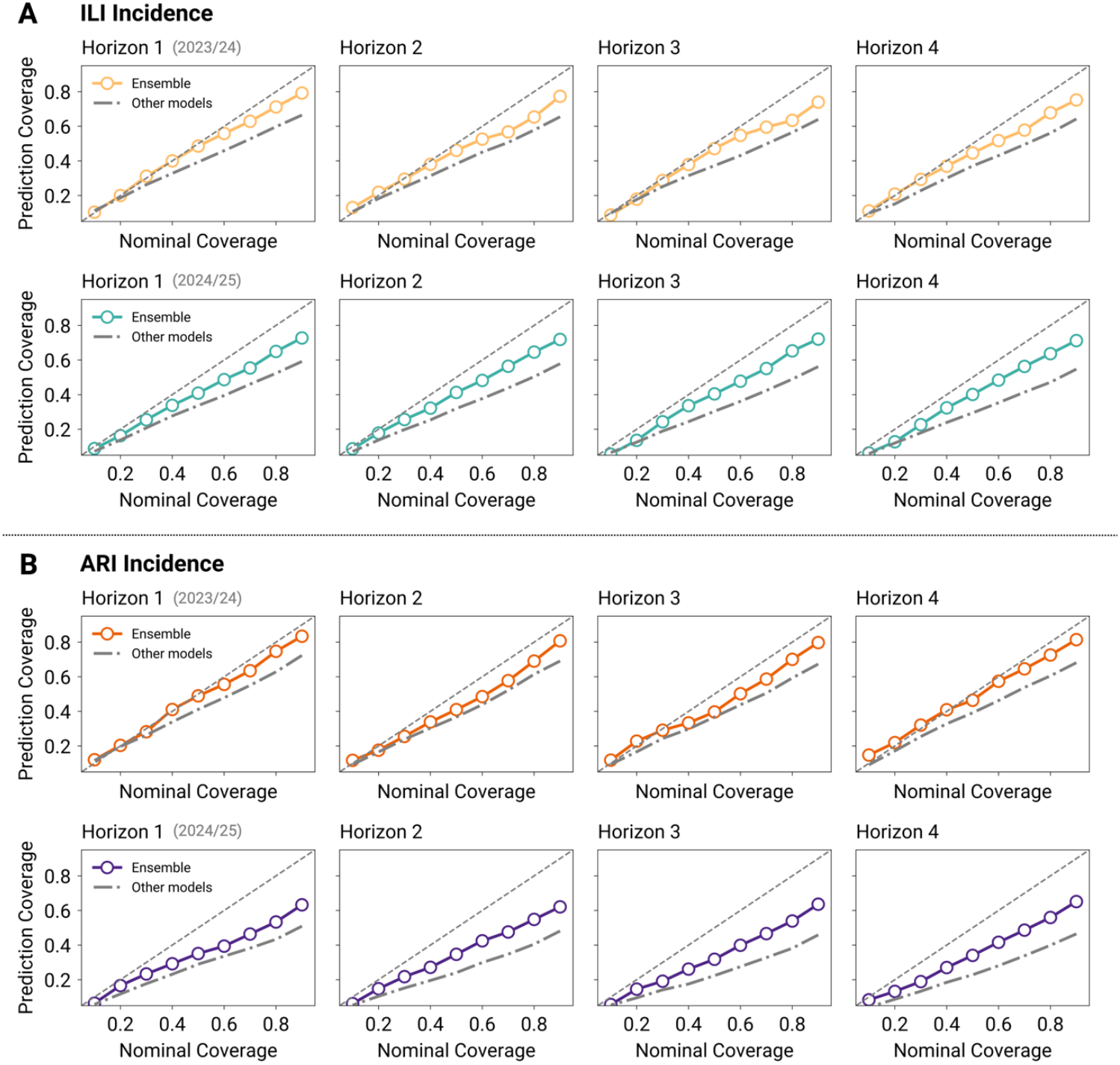
A) Nominal versus ensemble prediction coverage across forecasting horizons from 1 to 4 weeks ahead, for ILI (A) and ARI (B) incidence forecasting targets for the 2023/24 and 2024/25 forecasting seasons (top and bottom row of each subpanel, respectively). Results are shown for the ensemble (colored circles) and average of individual models (dash-dot line). Ideal calibration corresponds to the diagonal line.

In 2023/24, for ILI incidence, the ensemble forecasts align well with the diagonal across horizons at lower nominal coverage levels (i.e., up to 50%), indicating good overall calibration in that range. For higher nominal coverage levels, a consistent trend of overconfidence is observed, with the magnitude of undercoverage fairly constant with varying horizon length. For example, at 90% nominal coverage the prediction coverage is 79% at horizon 1 (−11% deviation from nominal coverage), and reduces to 75% at horizon 4 (−15% deviation). In 2024/25, we observe similar patterns, with overconfidence evident also at lower nominal coverage levels and overall larger deviations from nominal coverage. No clear trend is seen across increasing horizons. Nonetheless, in both seasons, the ensemble shows improved calibration across all levels and horizons compared to the pooled set of all other models.

ARI incidence exhibits a comparable pattern, with the ensemble forecasts showing improved calibration relative to individual models, and systematic undercoverage observed for all horizons at higher nominal coverage levels. Also for ARI incidence, coverage deviations are typically larger in 2024/25 than in 2023/24. No systematic degradation of calibration is observed as the forecast horizon increases. While the pooled analysis presented in the main text indicates overall reliable calibration for the ensemble, country-level results reveal heterogeneity. Supplementary Figure S21 summarizes deviations in 90% prediction coverage by country, highlighting substantial location-specific variability in ensemble forecast calibration.

## 4. Discussion

We evaluated the performance of ensemble forecasts in the context of the new ECDC forecasting hub, RespiCast, during the 2023/24 and 2024/25 winter seasons. The analysis shows the effectiveness of the ensemble in improving forecast accuracy and reliability compared to a naive baseline and individual models for ILI and ARI incidence across European countries. The ensemble outperformed the baseline model at most times and for most countries, and demonstrated greater reliability compared to individual models. The performance analysis also revealed challenges, mainly variation in forecasting performance over time and by country as well as overconfidence in the ensemble forecasts.

Forecasting performance (measured as relative WIS and AE compared to a naive baseline model) varied over time, with lower accuracy near the Christmas-New Year period in both seasons, followed by improvements as the season progressed. Several factors may have contributed to lower performance in these weeks. First, the Christmas-New Year period is a time when data reporting may be delayed and less accurate, potentially affecting model calibration and forecasting performance. Second, this period coincided with an increase or peak in ILI and/or ARI incidence in most countries, and thus represents one of the most challenging epidemic phases to predict. Third, fewer models were submitted during these rounds (as shown in Figure S1), especially in season 2023/24, potentially reducing the ensemble’s performance, which has been shown to be closely tied to the number of contributing models [18]. Finally, we note that the 2023/24 and 2024/25 winter seasons marked RespiCast’s inaugural seasons, and its performance may have been affected by a learning curve as new teams onboarded and refined their submissions. Further multi-season evaluation will be needed to disentangle the individual contributions of these factors to ensemble performance. It is important to acknowledge that the performance metrics used in this analysis are defined relative to a baseline model. It follows that their interpretation may be influenced by the baseline’s behaviour as well as the characteristics of the underlying data. For instance, during early phases and near the peak (i.e, where the incidence curve is flat), it is more challenging to improve over a persistence model, leading to low relative performances possibly assosciated with *good* absolute performances. This effect may be particularly relevant in countries where little or no improvement over the baseline was observed: in settings with very flat reported incidence time series, a persistence baseline is already hard to beat, so low relative performance can coexist with an ensemble that is nevertheless tracking the data well in absolute terms.

RespiCast provided forecasts for 26 European countries, revealing substantial variability in performance across locations. This indicates the potential impact on forecast accuracy of different local epidemic dynamics or differences in surveillance systems. Identifying the specific country-level factors that drive these differences remains challenging without extensive local knowledge and forecasts from several seasons. Additionally, the option for modelling teams to selectively submit forecasts across countries and rounds may introduce bias, as models might opt out of settings where they previously underperformed, potentially skewing ensemble reliability in some locations. In the future, when additional seasons of forecasts become available, we may be able to further investigate the underlying drivers of this heterogeneous performance between locations. Forecast accuracy is also inherently tied to the availability and quality of underlying data. Differences in data completeness, timeliness, and consistency across countries can significantly affect model calibration and evaluation. For instance, irregular updates or retrospective revisions in surveillance data may distort accuracy of forecasts or limit their real-time utility. These challenges underscore the need for continued investment in harmonized and high-quality surveillance systems across Europe.

Future analyses will be needed to examine how model participation and methodological heterogeneity influence RespiCast ensemble performance. In this context, a key open question is how to optimize ensemble composition. Understanding the role of model type, quality, and complementarity in shaping ensemble effectiveness could inform strategies for model selection and weighting aimed, for instance, at improving ensemble calibration and addressing the systematic overconfidence observed. In addition, active management of contributions, by promoting participation during critical periods and encouraging methodological diversity, may further improve ensemble performance, ultimately increasing utility of forecasts for public health decision-making.

An important methodological consideration concerns the definition of relative performance metrics. In our analysis, we calculated relative scores on a per-round basis (i.e., computing the ratio of baseline to model WIS for each forecasting round separately before aggregating across rounds for visualization). This approach provides intuitive insights into spatiotemporal performance patterns and facilitates, for example, the identification of periods and countries with distinct forecasting challenges. It is also particularly valuable given the substantial heterogeneity observed in forecasting performance across epidemic phases. Nonetheless, alternative definitions of relative performance exist. To assess the robustness of our findings to this methodological choice, we conducted a sensitivity analysis (Supplementary Figure S22 and Table S3) computing a relative performance metric analogous to the one used in Ref. [6]. The results confirm our main conclusions and indicate that the observed performance advantages of the ensemble are robust and does not depend on the relative score definition.

While ILI and ARI incidence are widely reported syndromic indicators that enable consistent pan-European forecasting, they also present limitations as neither are pathogen-specific. Forecasting pathogen-specific targets may be more directly linked to transmission dynamics, leading to increased conceptual clarity. For example, in the future syndromic indicators could be multiplied by positivity rates to serve as pathogen-specific incidence proxies, similar to the Goldstein Index frequently used to gauge influenza activity [22]. This pathogen-specific approach could also support the evolution of RespiCast into a more integrated forecasting hub aligned with the multi-pathogen surveillance data reported within ERVISS, enhancing its value for situational awareness and public health decision-making.

Finally, further work will be needed to align forecasting efforts with public health needs. This can include, for example, extending evaluation metrics to be more relevant for decision-makers and improving forecasting performance during critical epidemic phases. A user-focused assessment could guide next steps and help tailor future iterations of RespiCast to better support public health surveillance and response.

In conclusion, the RespiCast ensemble model outperformed the baseline and individual models during both 2023/24 and 2024/25 winter season in terms of WIS and absolute error across different countries and forecasting rounds. These seasons served as a valuable proof of concept, identifying key areas for improvement, such as overconfidence and performance variability across times and locations. RespiCast establishes a solid foundation for advancing forecasting capabilities in Europe and supporting public health decision-making.

## Data Availability

All the data used in this manuscript are available online at:

- 2023/24:
  - ILI: https://github.com/european-modelling-hubs/flu-forecast-hub_archive
  - ARI: https://github.com/european-modelling-hubs/ari-forecast-hub_archive
- 2024/25: https://github.com/european-modelling-hubs/RespiCast-SyndromicIndicators

## Acknowledgements and Funding

Data acknowledgment: ERVI-Net (European Respiratory Virus Surveillance Network) reporting data to TESSy.

Funding acknowledgment: This work received funding from the European Centre for Disease Prevention and Control (ECDC) via contract ECDC/2023/045 LOT1. V.M. acknowledges full and M.M partial support from the Lagrange Project of the ISI Foundation, funded by Fondazione CRT. K.S. is supported by Wellcome Trust (210758/Z/18/Z). A.A. is supported by the Luxembourg Government through the CoVaLux programme (16954531). D.PR. and G.G. are funded by the European Union through the ERC INSPIRE grant (project number 101076926). Views and opinions expressed are however those of the authors only and do not necessarily reflect those of the European Union or the European Research Council Executive Agency. Neither the European Union nor the European Research Council Executive Agency can be held responsible for them. Y.L. acknowledges support from the China Scholarship Council (CSC). K.E.S. and T.A.P. received support from the NIH National Institute of General Medical Sciences R35 MIRA program (grant no. R35GM143029). D.K. and F.M. were funded by the Ministry of Health, Welfare and Sport (VWS) in the Netherlands. F.M. also acknowledges funding from the Japan Society for the Promotion of Science (JSPS KAKENHI, 20J00793) and JST (JPMJPR23RA).

Other acknowledgments: Y.L. and N.P. thank the High-Performance Computing facilities at Queen Mary University of London. N.G. and E.B. thank Sebastian Funk and Johannes Bracher for the useful discussion.

