## Supplementary Information for "Performance evaluation of RespiCast ensemble forecasts for primary care syndromic indicators of viral respiratory disease in Europe"

Nicolò Gozzi[1,*], Corrado Gioannini[1], Paolo Milano[1], Ivan Vismara[1], Luca Rossi[1], Marco Quaggiotto[2,1], Valeria Marras[1], Stefania Fiandrino[1,3], Mattia Mazzoli[1], Daniela Paolotti[1], Alessandro Vespignani[4,1], Francesco Celino[5], Lorenzo Zino[5], Alessandro Rizzo[5], Sasikiran Kandula[6], Birgitte Freiesleben de Blasio[6,7], Maikel Bosschaert[8,9], Steven Abrams[8,10], Niel Hens[8,11], Atte Aalto[12], Daniele Proverbio[13], Giulia Giordano[13], Jorge Goncalves[12,14], Katharine Sherratt[15], Rhys Earl[16], Kelsey E. Shaw[17], T. Alex Perkins[17], Yuhan Li[18], Nicola Perra[18,19], Fuminari Miura[20,21,22], Don Klinkenberg[20,23], Rok Grah[24], Helen Johnson[24], Ajibola Omokanye[25], Leah J. Martin[25], Rene Niehus[25], Jose Canevari[25], Eva Bons[25]

[1]ISI Foundation, Turin, Italy
[2]Department of Design, Politecnico di Milano
[3]Department of Computer, Control, and Management Engineering Antonio Ruberti, Sapienza University of Rome, Rome, Italy
[4]Laboratory for the Modeling of Biological and Socio-technical Systems, Northeastern University, Boston, MA USA
[5]Department of Electronics and Telecommunications, Politecnico di Torino, Turin, Italy
[6]Norwegian Institute of Public Health, Oslo, Norway
[7]Department of Biostatistics, Institute of Basic Medical Sciences, University of Oslo
[8]Interuniversity Institute for Biostatistics and statistical Bioinformatics, Data Science Institute, UHasselt, Diepenbeek, Belgium
[9]Julius Center for Health Sciences and Primary Care University Medical Center Utrecht, Utrecht University
[10]Global Health Institute, Department of Family Medicine and Population Health, University of Antwerp, Antwerp, Belgium
[11]Centre for Health Economics Research and Modelling Infectious Diseases, Vaccine and Infectious Disease Institute, Antwerp, Belgium
[12]Luxembourg Centre for Systems Biomedicine, University of Luxembourg
[13]Department of Industrial Engineering, University of Trento, via Sommarive 9, Trento 38123, Italy
[14]Department of Plant Sciences, University of Cambridge
[15]Centre for Mathematical Modelling of Infectious Diseases, London School of Hygiene & Tropical Medicine, London, United Kingdom
[16]MRC Centre for Global Infectious Disease Analysis, Imperial College London
[17]Department of Biological Sciences, University of Notre Dame, Notre Dame, Indiana, USA
[18]School of Mathematical Sciences, Queen Mary University of London, UK
[19]The Alan Turing Institute, London, UK
[20]Centre for Infectious Disease Control, National Institute for Public Health and the Environment (RIVM), Bilthoven, the Netherlands
[21]Center for Marine Environmental Studies, Ehime University, Ehime, Japan
[22]Institute of Tropical Medicine, Nagasaki University, Nagasaki, Japan
[23]Department of Animal Sciences, Wageningen University and Research, Wageningen, the Netherlands
[24]Safinea
[25]European Centre for Disease Prevention and Control

*

### S1. Submitting Models and Participation

In Table S1 we report the list of participating teams and models in season 2023/24 and 2024/25. A short description summarising methods and approach of different models is also reported. Table S2 reports, for each individual model, the number and percentage of rounds in which the model contributed at least one forecasts (i.e., one country) in each season. Finally, Figure S1 shows the number of individual models submitted in each forecasting round for both targets and seasons.

| **Submitting Models – Syndromic Indicators** | | | |
|---|---|---|---|
| **Team** | **Model Name** | **Description** | **Season** |
| CSL_PoliTo | *metaFlu* | An epidemic model based on the susceptible-exposed-infectious-noninfectious-removed dynamic, with meta-population and activity-driven modelling, class subdivision, Bayesian parameter optimization | 2023/24 |
| ECDC | *FluForARIMA* | A simple ARIMA model with seasonality | 2023/24 |
| | *norrsken_blue* | A Bayesian piecewise square-root-linear model fit.<br><br>Model consists of 1) scaling of the data (convert to square-root-scale), 2) assigning data points into groups such that neighbouring data points belong to the same group, and simultaneously in Stan 3) fitting piecewise intercept-slope models in each group, 4) regularizing the difference between neighbouring slopes to be close to 0, 5) generating a future slope based on the last slope plus fitted noise. | 2023/24 |
| | *norrsken_green* | A Bayesian piecewise log-linear model fit.<br><br>Model consists of 1) scaling of the data (convert to log-scale), 2) assigning data points into groups such that neighbouring data points belong to the same group, and simultaneously in Stan 3) fitting piecewise intercept-slope models in each group, 4) regularizing the difference between neighbouring slopes to be close to 0, 5) generating a future slope based on the last slope plus fitted noise. | 2023/24 |
| | *soca_simplex* | Using historical data patterns with highest similarity to current data to forecast the future values.<br><br>Model consists of 1) taking 'm' latest data points, 2) find 'n' closest neighbours from all historical data using L2-norm, 3) use the next data time point from each historical data points, 4) use data from point 3 to fit a log-normal distribution , 5) use the log-normal distribution to estimate the quantiles/distribution, 6) return to step 3 but use two data points ahead for estimations of horizon 2, etc for horizon 3 and 4, 7) find optimal values of 'n' and 'm for a given country, using past 4 weeks of forecasts. | 2023/24, 2024/25 |
| | *SARIMA* | A simple ARIMA model with seasonality | 2024/25 |
| | *ARI2MA* | An ARIMA model with seasonality and with sqrt transformed data | 2023/24 |
| | *ETS_AAN* | ETS model Additive error and trend, no seasonality. | 2023/24 |
| | *ETS_Auto* | ETS model where R automatically selects the best ETS model (error, trend, seasonality) based on the Akaike Information Criterion (AIC) or the Bayesian Information Criterion (BIC). | 2023/24 |

| | | | |
|---|---|---|---|
| ISI | *EpiNow* | A semimechanistic model based on Rt estimation developed by the epiforecasts team at LSHTM | 2023/24 |
| | *FluABCaster* | A stochastic, age structured compartmental model calibrated via ABC-SMC techniques. | 2023/24, 2024/25 |
| | *GLEAM* | A stochastic, age-structured compartmental model based on a metapopulation approach that uses real-world data on populations and human mobility to simulate epidemic spreading on a global scale. | 2023/24, 2024/25 |
| | *GenLogModel* | A generalized logistic growth model. | 2023/24 |
| | *IPSICast* | An exogenous autoregressive model that integrates information on digital surveillance data from the Influweb participatory system. | 2023/24 |
| | *FluBcast* | A stochastic, age-structured compartmental model that explicitly integrates behavioral changes through a new compartment of susceptible individuals who are risk averse. | 2024/25 |
| | *LSTFlu* | Bidirectional Long Short Term Memory model trained over 10 years of ILI surveillance data from Italy. | 2024/25 |
| | *SEIR-BRW* | SEIR Beta Random Walks is a stochastic compartmental model with random variations in the transmission rate, that provides short-term incidence forecasts. | 2024/25 |
| | *RC_AdaptEns2* | An adaptive ensemble of multiple models contributing to RespiCompass scenario projections for ILI. | 2024/25 |
| | *AriABCaster* | A stochastic, age structured compartmental model calibrated via ABC-SMC techniques. | 2023/24 |
| | *EpiNowARI* | A semimechanistic model based on Rt estimation developed by the epiforecasts team at LSHTM (https://epiforecasts.io/EpiNow2/) | 2023/24 |
| | *GenLogModelAri* | A generalized logistic growth model for ARI. | 2023/24 |
| ItaLuxColab | *EpiEKF* | Stochastic SIRS model with automatic data integration by the extended Kalman filter (EKF) with adaptive hyperparameter estimation, applied separately on each region (github.com/AtteAalto/EpiEKF) | 2023/24, 2024/25 |
| | *EpiNetEKF* | Stochastic networked SIRS model with automatic data integration by the extended Kalman filter (EKF) and adaptive hyperparameter estimation (gitlab.com/uniluxembourg/lcsb/systems-control/epinetekf) | 2024/25 |
| QMUL | *ARIMA* | Autoregressive integrated moving average model. | 2023/24, 2024/25 |
| | *SEIR* | Stochastic age-structured SEIR model. | 2023/24, 2024/25 |
| | *SEIR-augmented* | An SEIR compartmental model augmented by incorporating ARIMA model predictions for improved forecast selection | 2024/25 |
| fjordhest | *ensemble* | An inverse-WIS weighted ensemble of 3 component models - an exponential trend smoothing (ETS) model, a quantile AR model, and a baseline model of random walk with drift.<br><br>Quantile autoregression model uses quantgen R package, and is similar to CMU-Timeseries model from 2022/23 FluSight season. | 2023/24, 2024/25 |

| | | | |
|---|---|---|---|
| | | Random walk and ETS models use fable R package. To build an ensemble, the quantile distributions of the component models are weighted (location- and target-specific) by the mean of inverse-WIS scores over the last 3 weeks that could be evaluated. The estimates are solely the responsibility of the contributors and do not represent, nor are endorsed by, the Norwegian Institute of Public Health. | |
| MRC_GIDA | *NBEATS* | Neural Basis Expansion Analysis for Time Series | 2024/25 |
| | *CATBoost* | Categorical Boosting model | 2024/25 |
| | *NHiTS* | Neural Hierarchical Interpolation for Time Series | 2024/25 |
| | *Prophet* | Facebook Prophet forecasting model | 2024/25 |
| | *TiDE* | Time Series Dense Encoder | 2024/25 |
| NotreDame | *Rt_cocirc* | Smoothed ILI historic data used to estimate the Rt using the EpiEstim package in R. Rt then predicted into the future by a GAM model with smoothed COVID and Influenza data as predictors | 2024/25 |
| Safinea | *syndromic* | A state-space model with respiratory disease as a hidden states and separate observation processes for ILI and ARI reporting. Smoothed with unscented Kalman Filter, poisson process | 2024/25 |
| | *PRFM* | The PRFM model uses historical time-series motifs to detect similarities between the most recent data and past observations, and using principles from deterministic chaos theory to forecast the future incidence based on assigning weights to historical data and combining those to an ensemble-style forecast. Importantly, its hyperparameter tuning is driven by maximizing RespiCast top rank rather than minimizing WIS, avoiding issues related to combining past low and high WIS scores for optimization. | 2024/25 |
| RIVM | *KFdlm* | A dynamic linear model with the local trend and the first and second derivative terms, using a Kalman filter implemented in R-package dlm with log(x+1) transformation of the time series. | 2024/25 |

**Table S1**. Participating teams and models in the 2023/24 and 2024/25 RespiCast forecasting season.

| Model | 2023/24 | | 2024/25 | |
|---|---|---|---|---|
| | ILI incidence | ARI incidence | ILI incidence | ARI incidence |
| *ARI2MA* | / | 20 (100.0%) | / | / |
| *ARIMA* | 15 (75.0%) | / | 26 (92.9%) | / |
| *AriABCaster* | / | 20 (100.0%) | / | / |
| *CATBoost* | / | / | 1 (3.6%) | 1 (3.6%) |
| *ETS_AAN* | / | 1 (5.0%) | / | / |
| *ETS_Auto* | / | 1 (5.0%) | / | / |
| *EpiEKF* | 12 (60.0%) | / | 26 (92.9%) | / |
| *EpiNetEKF* | / | / | 26 (92.9%) | / |
| *EpiNow* | 13 (65.0%) | / | / | / |
| *EpiNowARI* | / | 13 (65.0%) | / | / |
| *FluABCaster* | 20 (100.0%) | / | 23 (82.1%) | 22 (78.6%) |
| *FluBcast* | / | / | 28 (100.0%) | 28 (100.0%) |
| *FluForARIMA* | 20 (100.0%) | / | / | / |
| *GLEAM* | 15 (75.0%) | / | 24 (85.7%) | 24 (85.7%) |
| *GenLogModel* | 11 (55.0%) | / | / | / |
| *GenLogModelAri* | / | 20 (100.0%) | / | / |
| *IPSICast* | 14 (70.0%) | / | / | / |
| *KFdlm* | / | / | 26 (92.9%) | / |
| *LSTFlu* | / | / | 22 (78.6%) | 21 (75.0%) |
| *NBEATS* | / | / | 1 (3.6%) | 1 (3.6%) |
| *NHiTS* | / | / | 1 (3.6%) | 1 (3.6%) |
| *Prophet* | / | / | 1 (3.6%) | 1 (3.6%) |
| *RC_AdaptEns2* | / | / | 28 (100.0%) | / |
| *Rt_cocirc* | / | / | 23 (82.1%) | 21 (75.0%) |
| *SARIMA* | / | / | 27 (96.4%) | 27 (96.4%) |
| *SEIR* | 18 (90.0%) | / | 23 (82.1%) | / |
| *SEIR_BRW* | / | / | 21 (75.0%) | 15 (53.6%) |
| *SEIR_agumented* | / | / | 23 (82.1%) | / |
| *ensemble* | 16 (80.0%) | 16 (80.0%) | 25 (89.3%) | 25 (89.3%) |
| *metaFlu* | 14 (70.0%) | / | / | / |
| *norrsken_blue* | 18 (90.0%) | 19 (95.0%) | / | / |
| *norrsken_green* | 18 (90.0%) | 19 (95.0%) | / | / |
| *soca_simplex* | 11 (55.0%) | 11 (55.0%) | 27 (96.4%) | 27 (96.4%) |
| *syndromic* | / | / | 11 (39.3%) | 11 (39.3%) |

**Table S2**. Number of participating forecasting rounds for each model contributing to ILI and ARI incidence targets in the 2023/24 and 2024/25 RespiCast seasons. The table reports both the absolute number of rounds and the corresponding percentage with respect to the total number of rounds in each season.

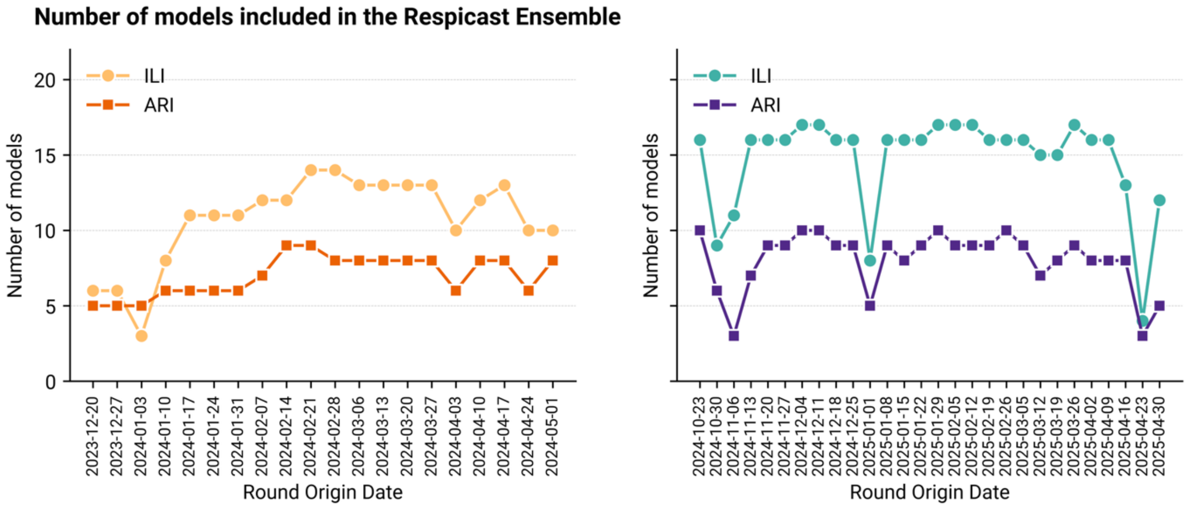


**Figure S1**. Number of models included in the RespiCast ensemble for ILI and ARI incidence forecasts across forecasting rounds during the 2023/24 (left) and 2024/25 (right) seasons. The x-axis represents the origin date of each forecasting round, while the y-axis shows the total number of distinct models submitted in each round.

### S2. Ensemble forecasts during 2023/24 and 2024/25 seasons

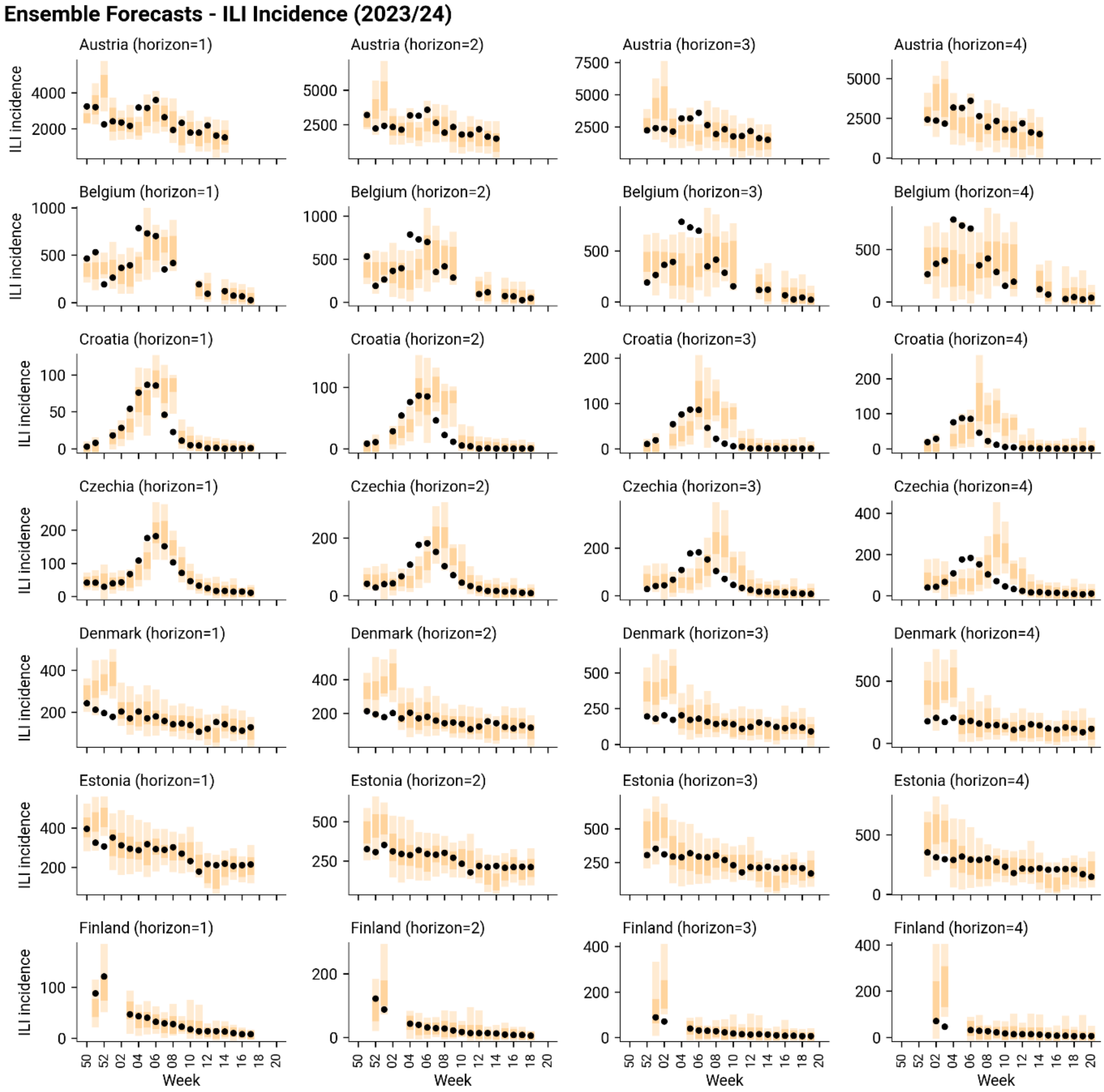


**Figure S2**. Ensemble forecasts of ILI incidence for Austria, Belgium, Croatia, Czechia, Denmark, Estonia, and Finland during the 2023/24 respiratory disease season. Each row shows forecasts at different horizons for a given country, with the x-axis indicating the epidemiological week and the y-axis showing ILI incidence. The yellow bars represent the ensemble forecast predictions, including 50% and 95% prediction intervals, while the black dots denote the reported data.

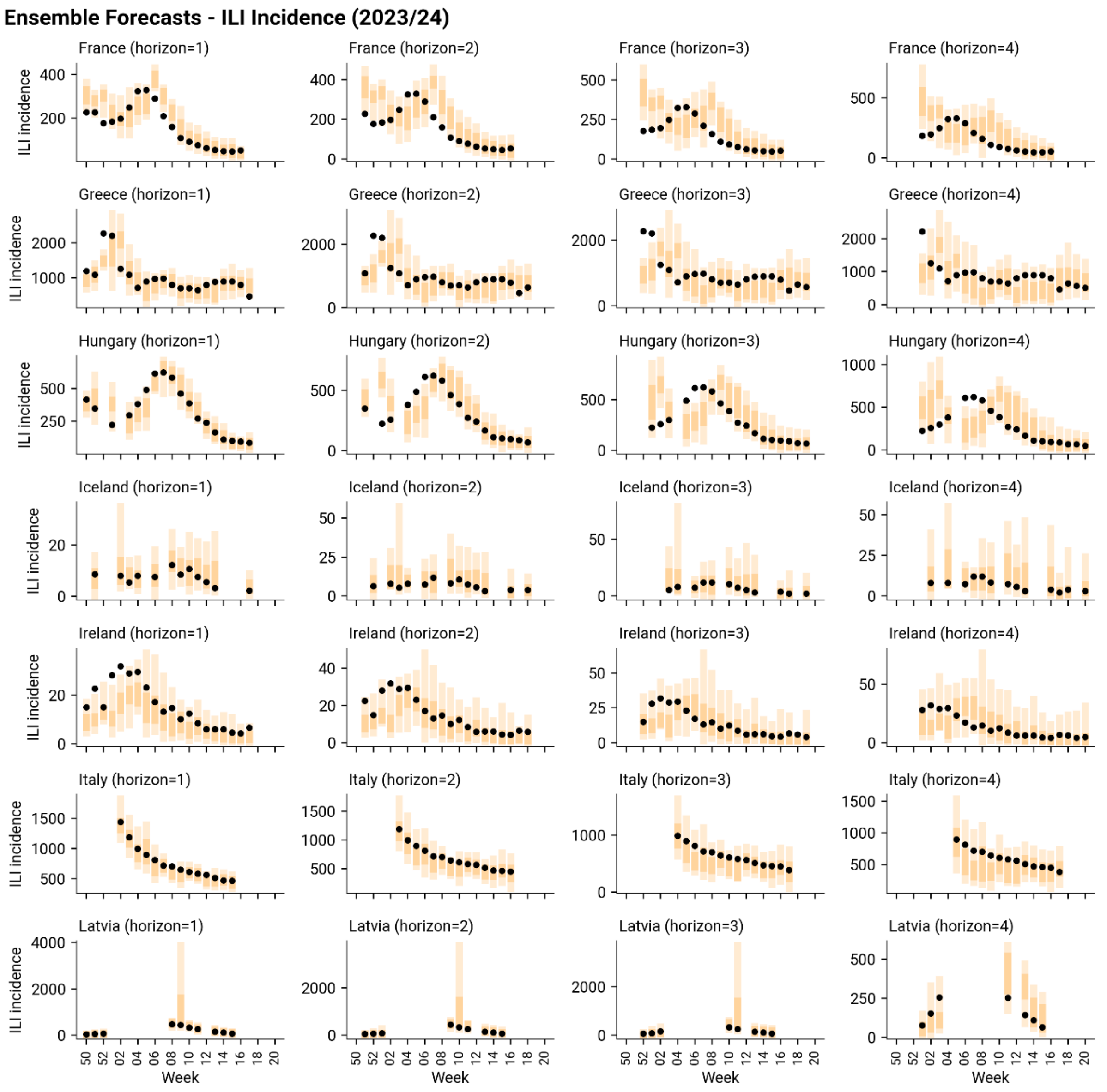


**Figure S3**. Ensemble forecasts of ILI incidence for France, Greece, Hungary, Iceland, Ireland, Italy, and Latvia during the 2023/24 respiratory disease season. Each row shows forecasts at different horizons for a given country, with the x-axis indicating the epidemiological week and the y-axis showing ILI incidence. The yellow bars represent the ensemble forecast predictions, including 50% and 95% prediction intervals, while the black dots denote the reported data.

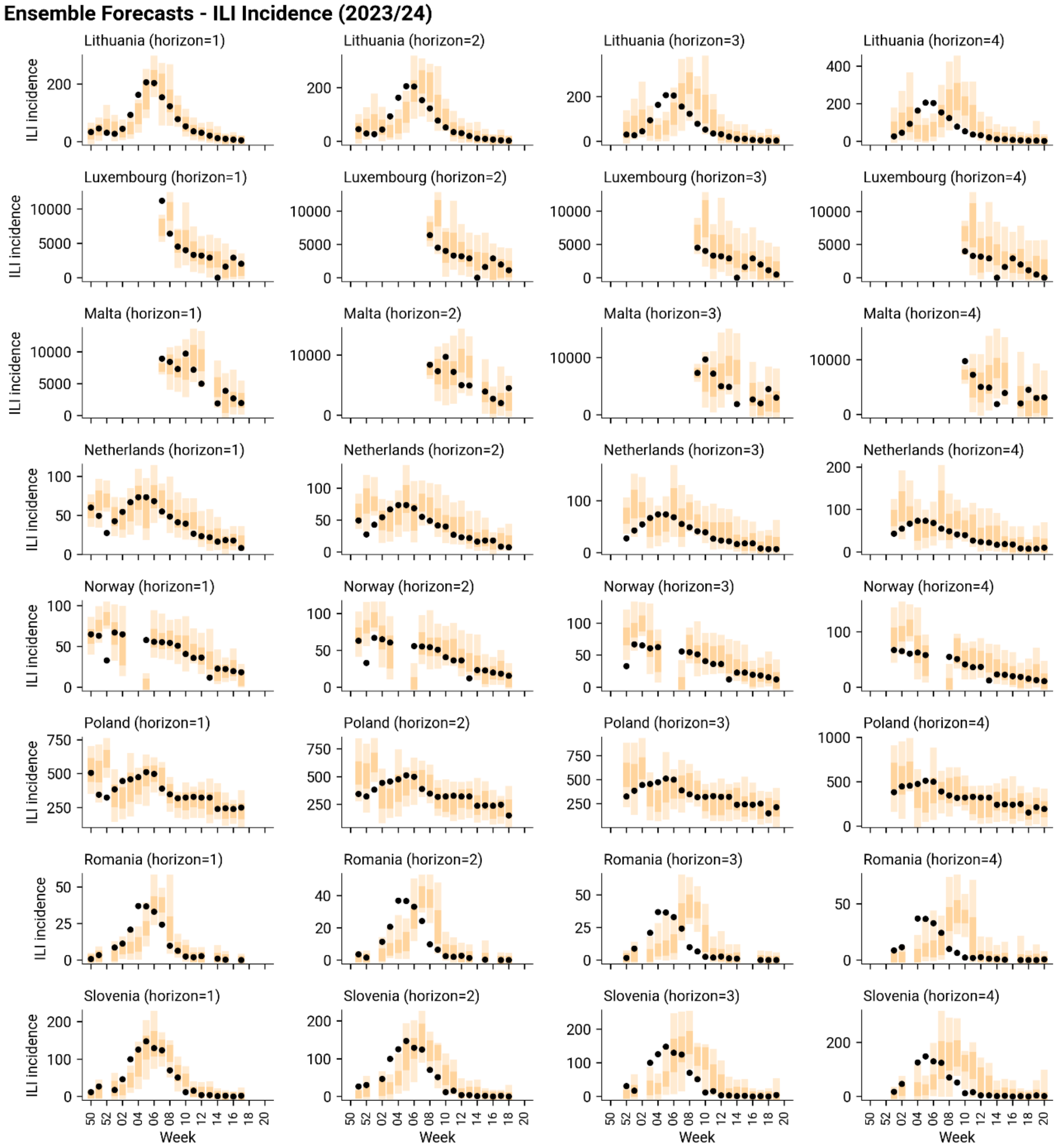


**Figure S4**. Ensemble forecasts of ILI incidence for Lithuania, Luxembourg, Malta, Netherlands, Norway, Poland, Romania, and Slovenia during the 2023/24 respiratory disease season. Each row shows forecasts at different horizons for a given country, with the x-axis indicating the epidemiological week and the y-axis showing ILI incidence. The yellow bars represent the ensemble forecast predictions, including 50% and 95% prediction intervals, while the black dots denote the reported data.

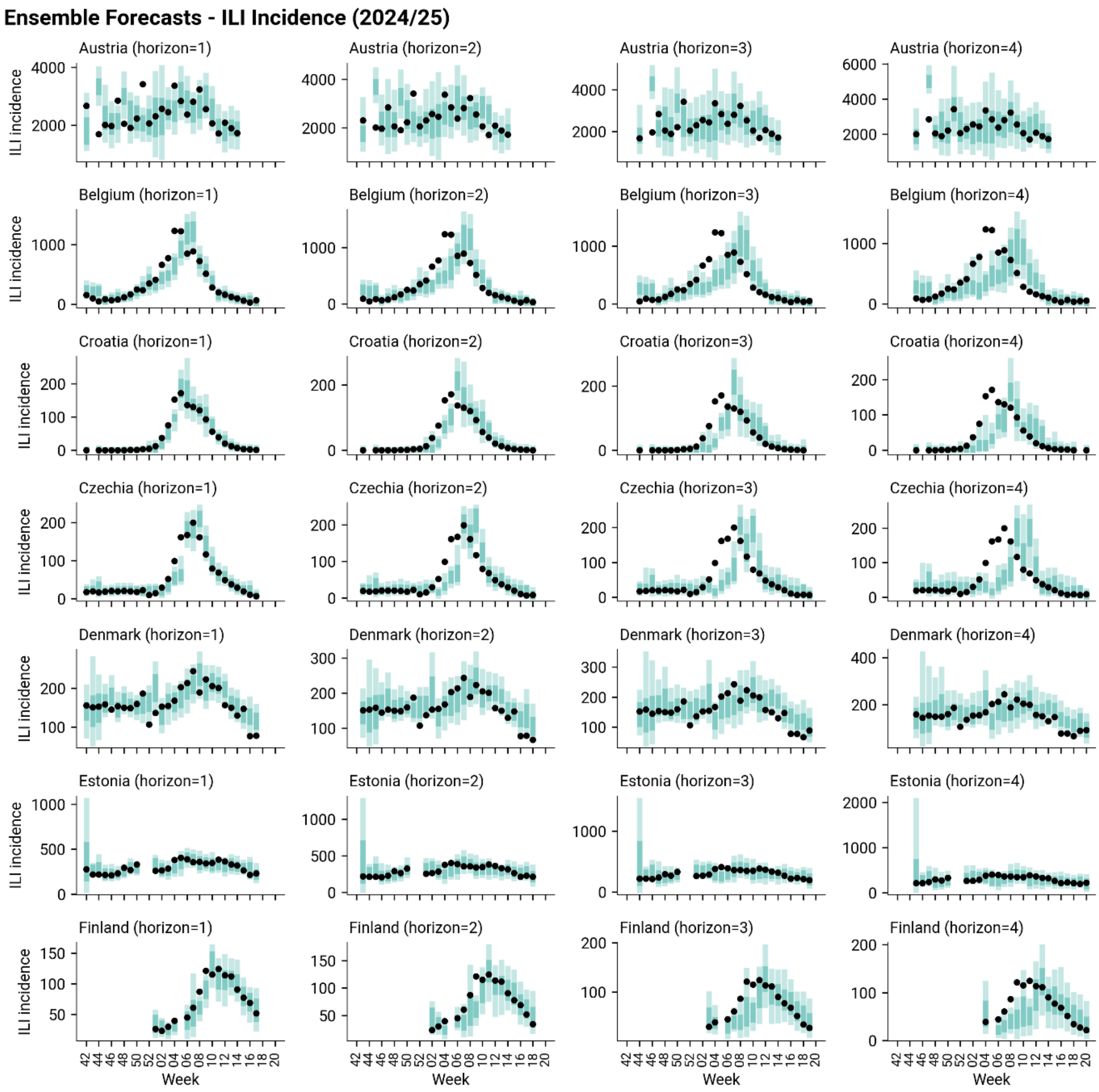


**Figure S5**. Ensemble forecasts of ILI incidence for Austria, Belgium, Croatia, Czechia, Denmark, Estonia, and Finland during the 2024/25 respiratory disease season. Each row shows forecasts at different horizons for a given country, with the x-axis indicating the epidemiological week and the y-axis showing ILI incidence. The blue bars represent the ensemble forecast predictions, including 50% and 95% prediction intervals, while the black dots denote the reported data.

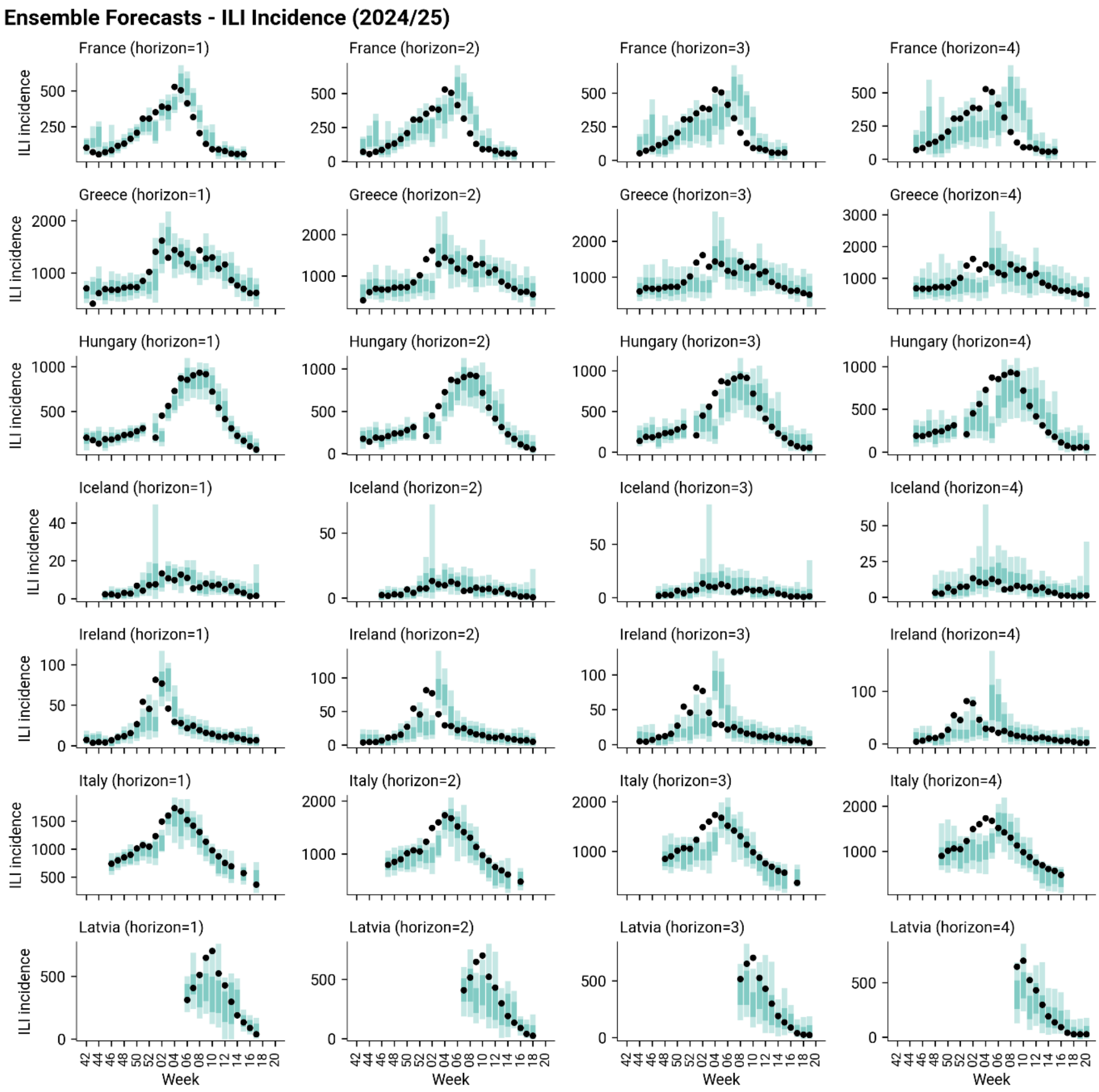


**Figure S6**. Ensemble forecasts of ILI incidence for France, Greece, Hungary, Iceland, Ireland, Italy, and Latvia during the 2024/25 respiratory disease season. Each row shows forecasts at different horizons for a given country, with the x-axis indicating the epidemiological week and the y-axis showing ILI incidence. The blue bars represent the ensemble forecast predictions, including 50% and 95% prediction intervals, while the black dots denote the reported data.

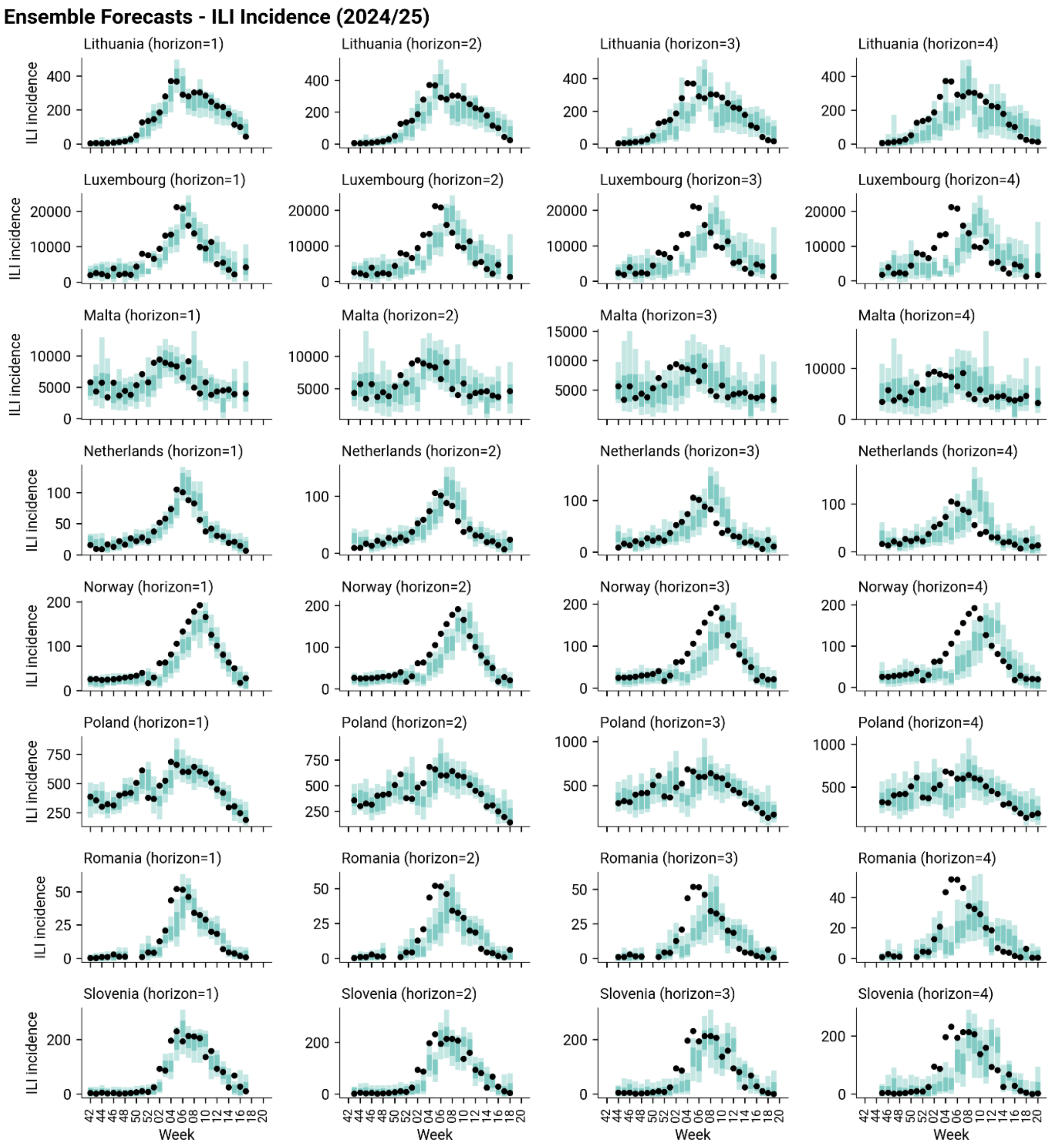


**Figure S7**. Ensemble forecasts of ILI incidence for Lithuania, Luxembourg, Malta, Netherlands, Norway, Poland, Romania, and Slovenia during the 2024/25 respiratory disease season. Each row shows forecasts at different horizons for a given country, with the x-axis indicating the epidemiological week and the y-axis showing ILI incidence. The blue bars represent the ensemble forecast predictions, including 50% and 95% prediction intervals, while the black dots denote the reported data.

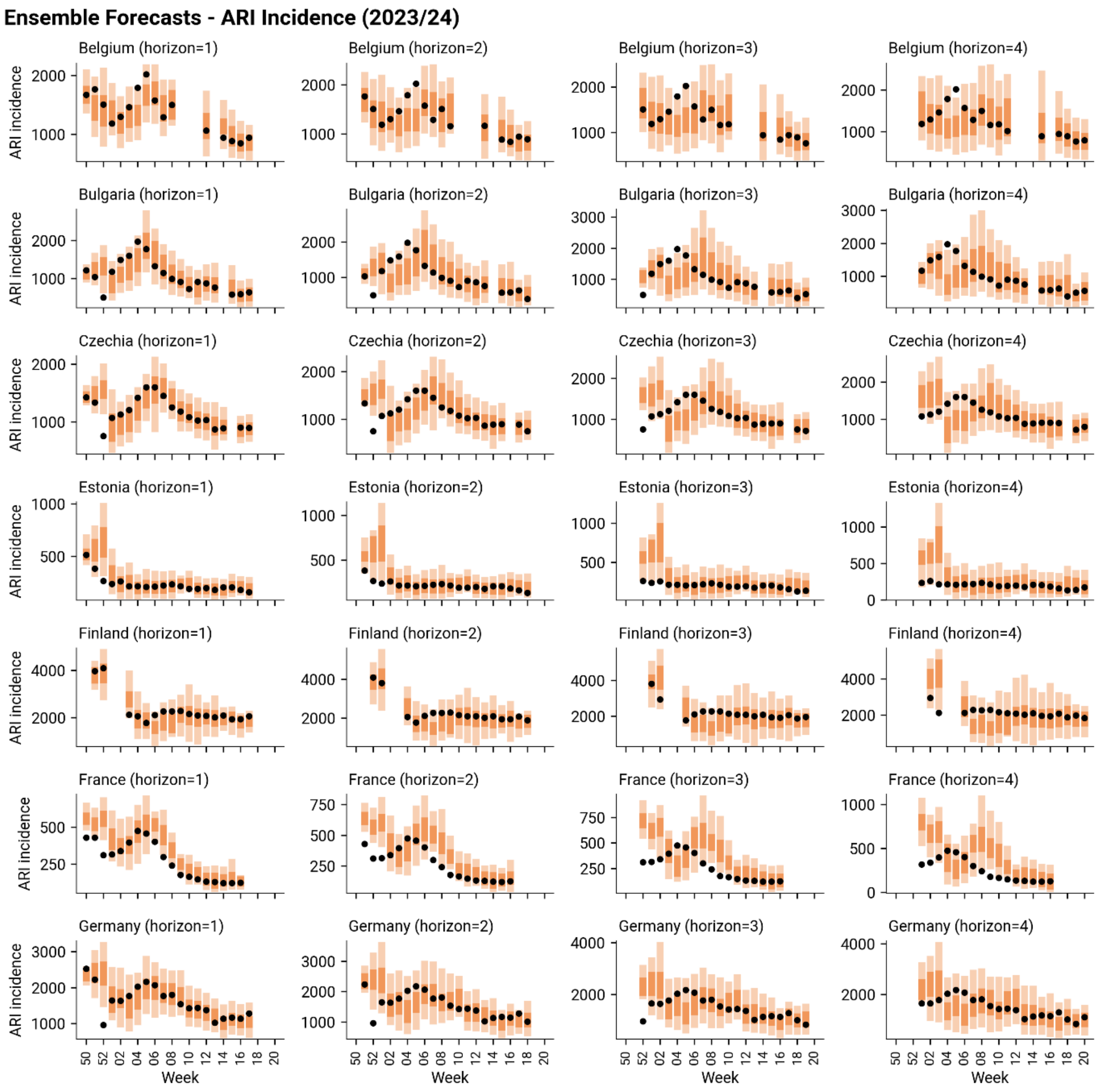


**Figure S8**. Ensemble forecasts of ARI incidence for Belgium, Bulgaria, Czechia, Estonia, Finland, France, and Germany during the 2023/24 respiratory disease season. Each row shows forecasts at different horizons for a given country, with the x-axis indicating the epidemiological week and the y-axis showing ILI incidence. The orange bars represent the ensemble forecast predictions, including 50% and 95% prediction intervals, while the black dots denote the reported data.

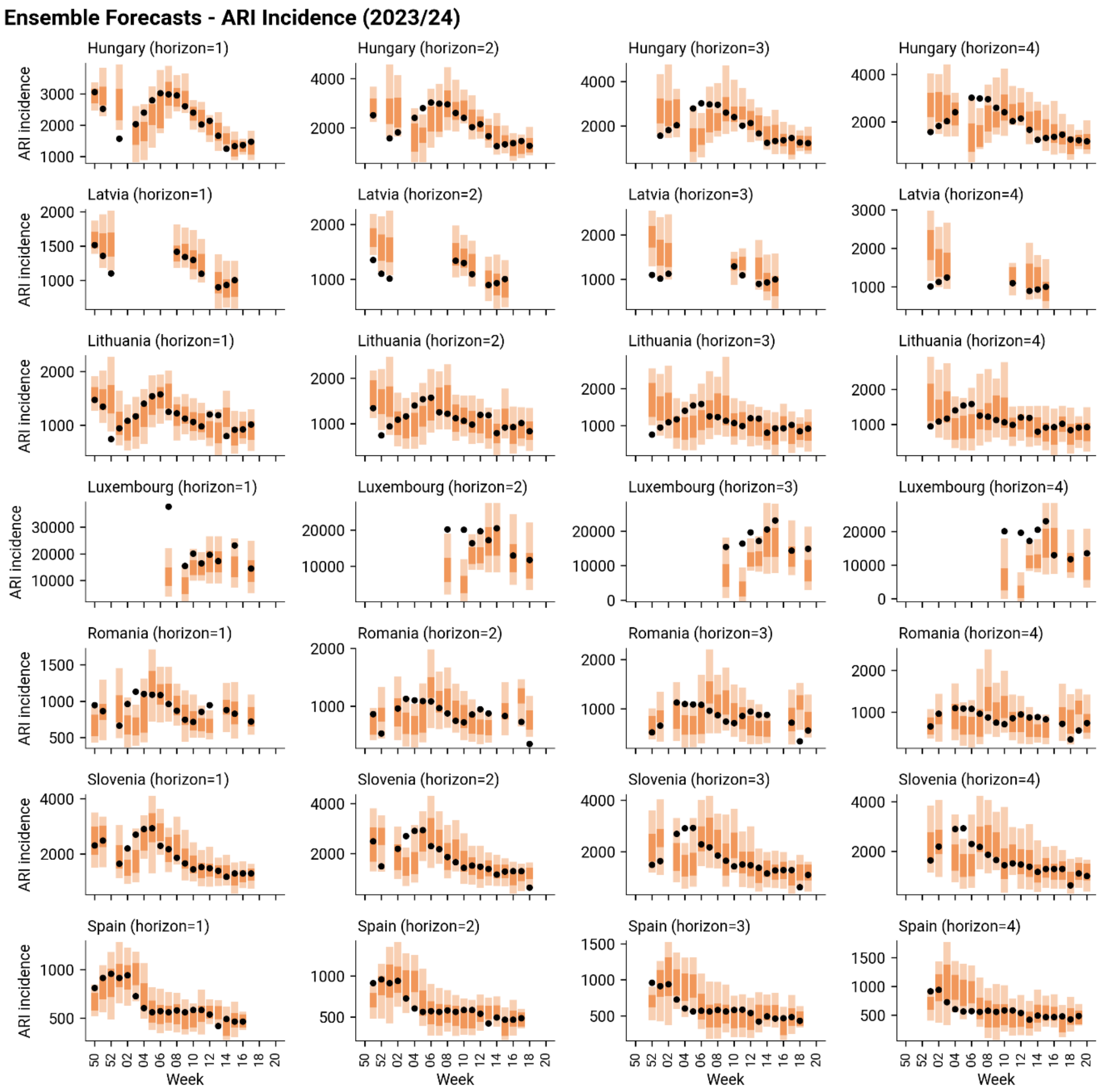


**Figure S9**. Ensemble forecasts of ARI incidence for Hungary, Latvia, Lithuania, Luxembourg, Romania, Slovenia, and Spain during the 2023/24 respiratory disease season. Each row shows forecasts at different horizons for a given country, with the x-axis indicating the epidemiological week and the y-axis showing ILI incidence. The orange bars represent the ensemble forecast predictions, including 50% and 95% prediction intervals, while the black dots denote the reported data.

#### Ensemble Forecasts - ARI Incidence (2024/25)

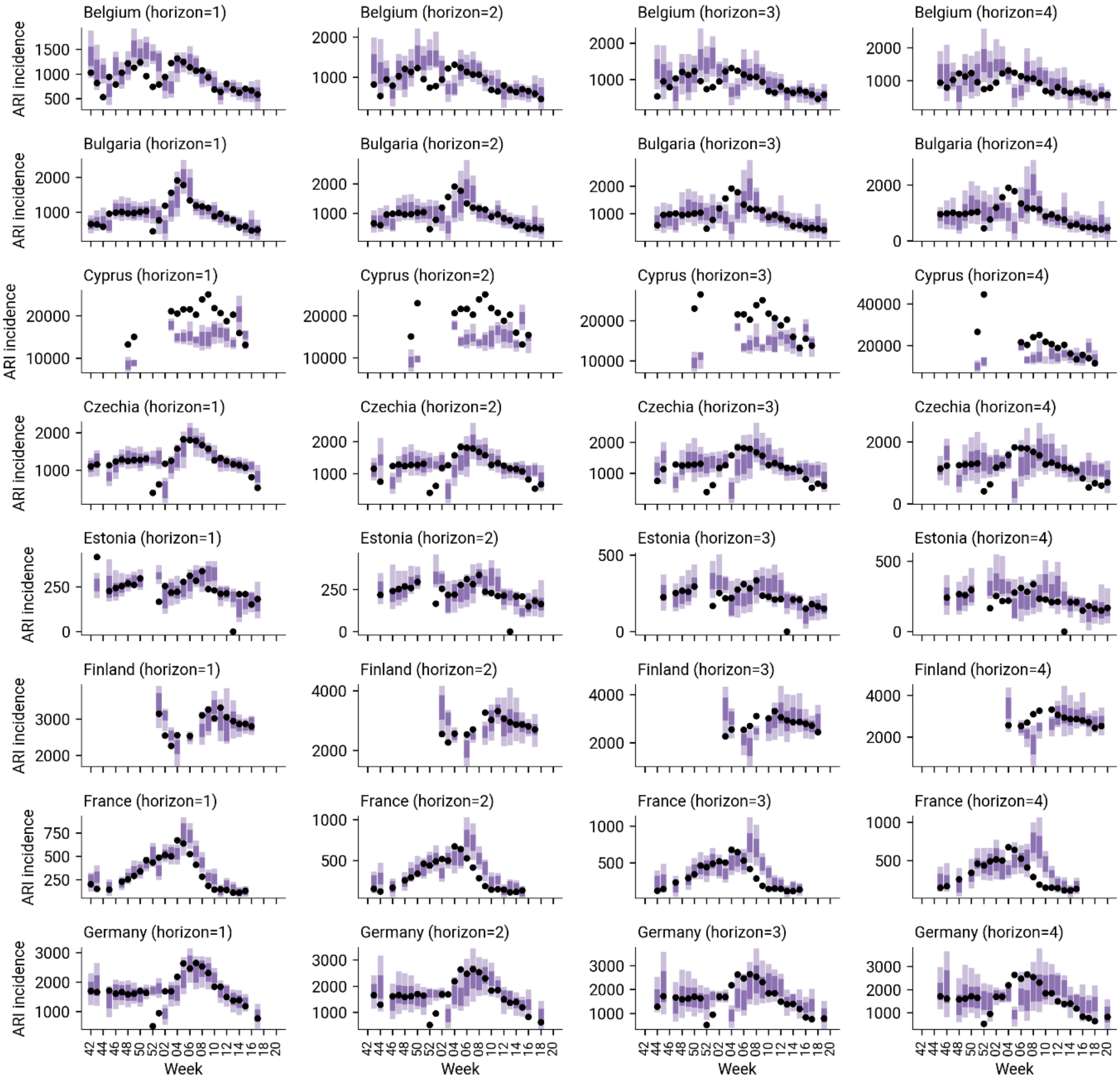


**Figure S10**. Ensemble forecasts of ARI incidence for Belgium, Bulgaria, Cyprus, Czechia, Estonia, Finland, France, and Germany during the 2024/25 respiratory disease season. Each row shows forecasts at different horizons for a given country, with the x-axis indicating the epidemiological week and the y-axis showing ILI incidence. The purple bars represent the ensemble forecast predictions, including 50% and 95% prediction intervals, while the black dots denote the reported data.

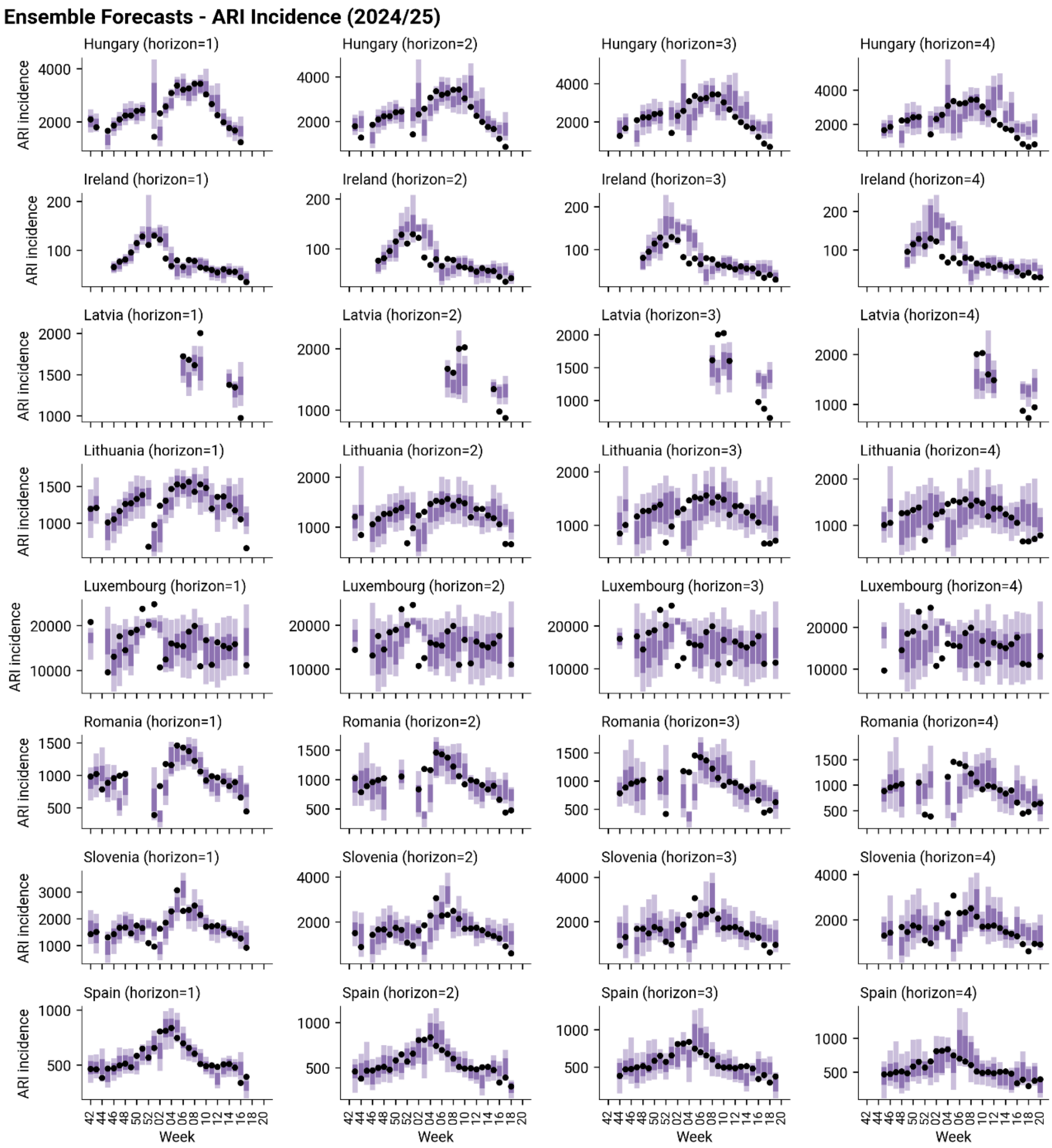


**Figure S11**. Ensemble forecasts of ARI incidence for Hungary, Ireland, Latvia, Lithuania, Luxembourg, Romania, Slovenia, and Spain during the 2024/25 respiratory disease season. Each row shows forecasts at different horizons for a given country, with the x-axis indicating the epidemiological week and the y-axis showing ILI incidence. The purple bars represent the ensemble forecast predictions, including 50% and 95% prediction intervals, while the black dots denote the reported data.

### S3. Forecasting performance during 2023/24 and 2024/25 seasons (Supplementary analyses)

#### S3.1 Relative AE performance

Figure S12 shows the performance of RespiCast ensemble forecasts for ILI incidence using the relative Absolute Error (AE) of the median. Panel A presents the distribution of relative AE values, aggregating results across all countries and forecasting rounds, separately for the ensemble and all other models combined. In season 2023/24, the median relative AE for individual models is 0.03, while the ensemble achieves a higher median relative AE of 0.24, demonstrating its superior predictive performance. Season 2024/25 shows similar patterns, with the ensemble achieving a median relative AE of 0.40, compared to 0.08 for all other individual models combined. Ensemble performance is also more stable, with an IQR of the relative AE distribution of 1.46 and 1.16 in the 2023/24 and 2024/25 seasons, respectively, compared to 1.86 and 1.56 for the distribution of other models combined. Panel B shows the temporal evolution of relative AE across epidemiological weeks in each season. Similar to relative WIS, the ensemble exhibits weaker performance in the early weeks of both seasons. Despite these initial challenges, it maintains a positive median relative AE in 80% (16/20) and nearly 90% (25/28) of weeks in seasons 2023/24 and 2024/25. Panel C displays the distribution of ensemble relative AE across countries, showing that the ensemble outperforms the baseline in 81% (18/22) and 100% (22/22) of countries in each season.  Panel D provides a granular view of relative AE by country and week, showing the average relative AE over the four-week forecasting horizon for forecasts produced in that week for each country. The observed performance patterns across locations and times align with those seen for relative WIS in the main text.

Figure S13 presents the same analysis for ARI incidence. Panel A shows that, in season 2023/24, individual models have a median relative AE of -0.19, while the ensemble's median relative AE is 0.10, indicating more modest improvements over the baseline. Similarly, in season 2024/25 the median relative AE of the ensemble is 0.11 compared to -0.17 of all the other individual models combined. Panel B confirms the initial lower performance in terms of AE, but in this case, challenges persist in later weeks, with 55% (11/20) and 75% (21/28) weeks showing a positive median relative AE in season 2023/24 and 2024/25, respectively. Panel C reveals that only 50% of the countries exhibit a positive median relative AE in 2023/24 (7/14), while this fraction increases to 87% (14/16) in 2024/25. Finally, Panel D reinforces previously observed patterns across locations and times while also highlighting persistent forecasting difficulties in some countries even in later weeks.

Overall, when considering relative AE, the improvements in forecasting performance of the ensemble compared to the baseline are smaller than those observed for relative WIS, suggesting that the magnitude of improvement varies by metric. Additionally, the analysis of relative AE further confirms that forecasting performance is generally better for ILI incidence compared to ARI incidence, with the ensemble showing stronger improvements over the baseline in the case of ILI. This is emphasized also in Figure S14 where we compare ensemble forecasts for ILI and ARI incidence both in terms of relative WIS and AE in each season.

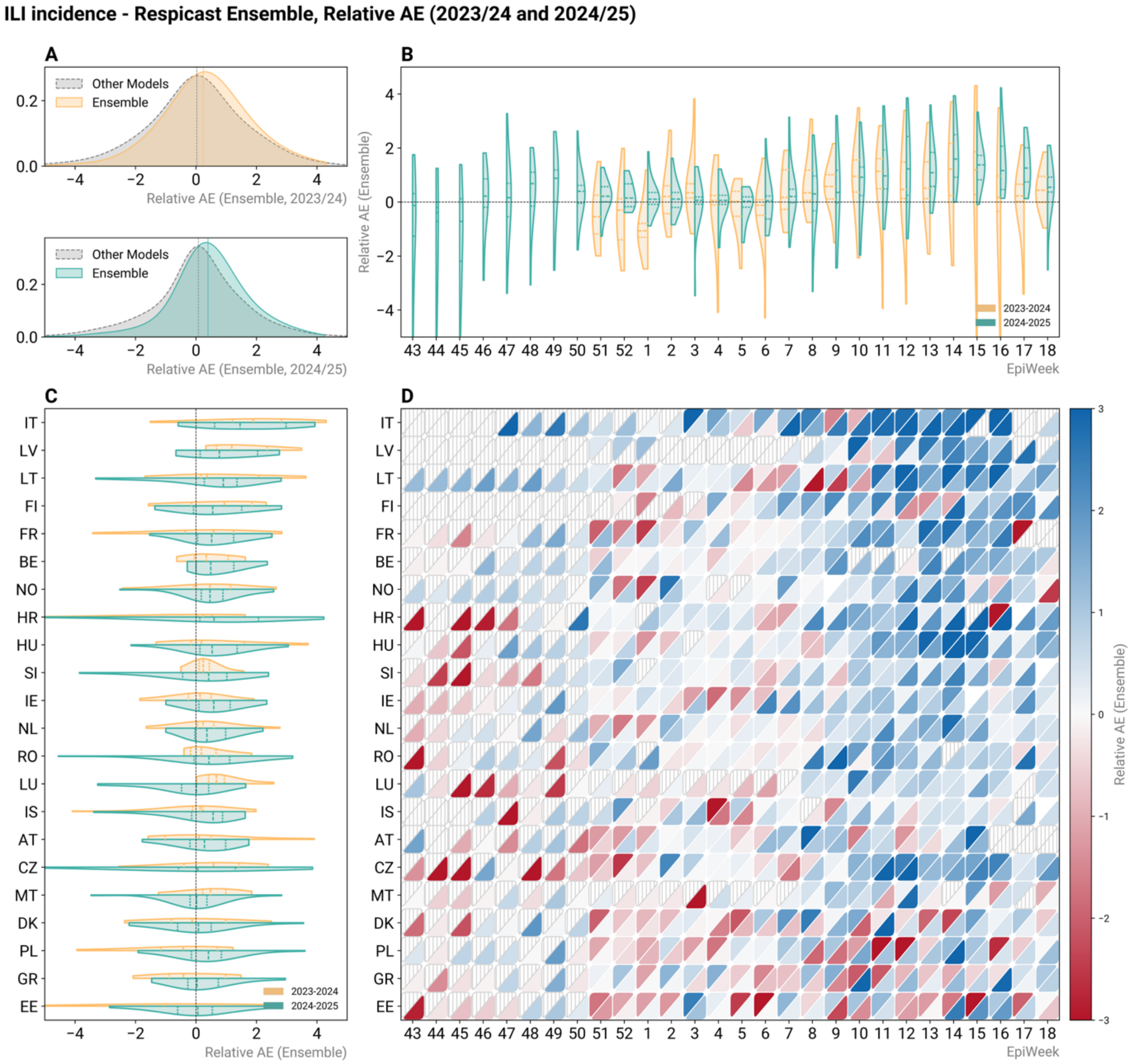


**Figure S12.** Relative Absolute Error (AE) of the median for Influenza-Like Illness (ILI) incidence predictions from the RespiCast Ensemble during the 2023/24 and 2024/25 winter seasons. **(A)** Distribution of relative AE across all countries and weeks, comparing the ensemble model to all other models combined in 2023/24 (top) and 2024/25 (bottom) winter seasons. **(B)** Distribution of relative AE of the ensemble across all countries for each epidemiological week separately (2023/24 in yellow, left half; 2024/25 in green, right half of the split violin). **(C)** Distribution of relative AE of the ensemble across all epidemiological weeks for each country separately (2023/24 in yellow, top half; 2024/25 in green, bottom half of the split violin). **(D)** Heatmap of relative AE values of the ensemble by country and epidemiological week, with blue indicating better performance (relative AE greater than 0) and red indicating worse performance (relative AE smaller than 0) compared to the baseline. The top-left half of each cell shows data for the 2023/24 season, while the bottom-right half shows data for 2024/25. Cells with grey stripes indicate missing data. The violin plots in Panels B and C are truncated at the observed minimum and maximum values, with dashed lines inside representing the 1st quartile, median, and 3rd quartile of the distribution. In all panels, relative AE values are averaged over the four-week forecasting horizon.

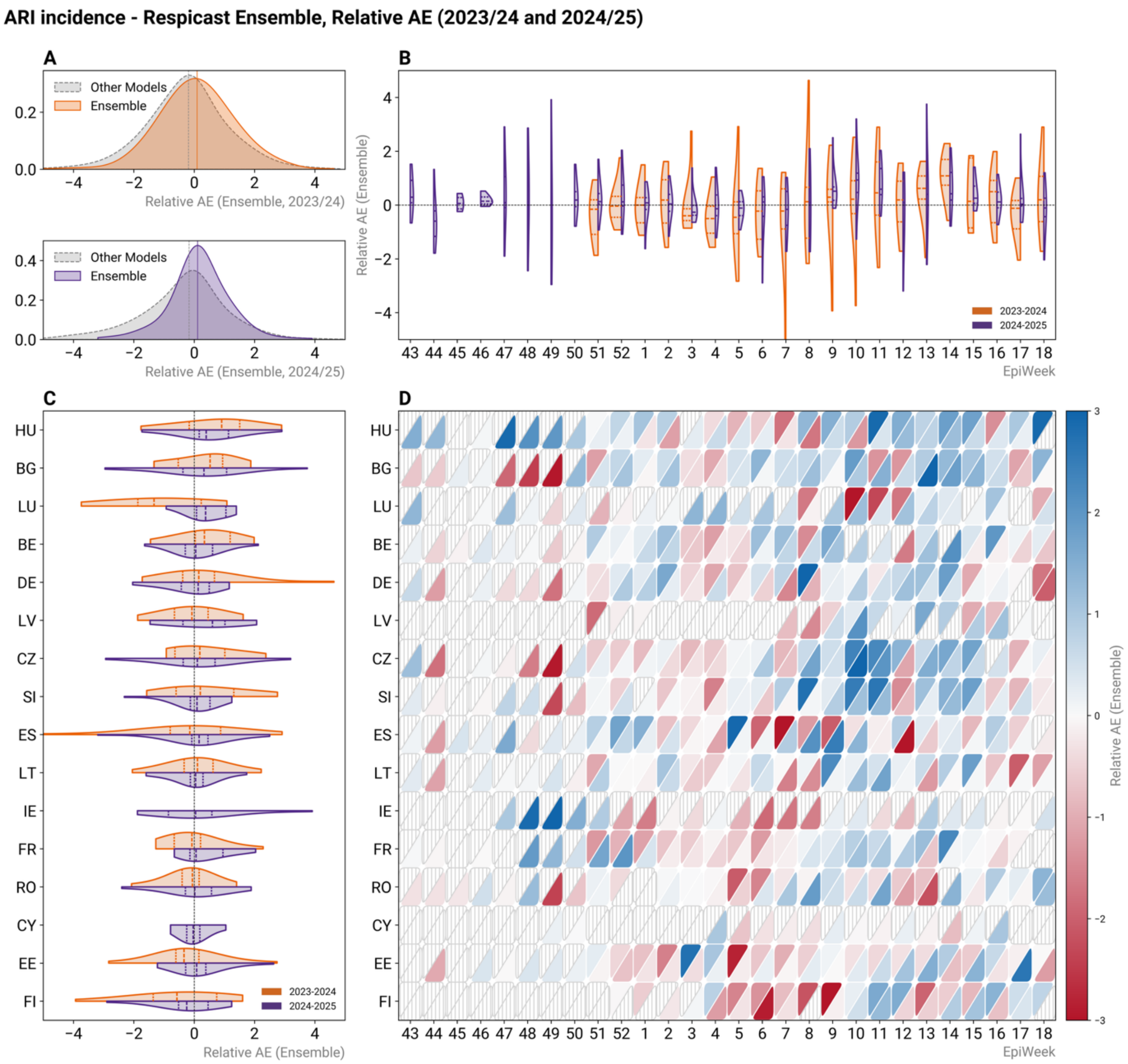


**Figure S13.** Relative Absolute Error (AE) of the median for Acute Respiratory Infection (ARI) incidence predictions from the RespiCast Ensemble during the 2023/24 and 2024/25 winter season. **(A)** Distribution of relative AE across all countries and weeks, comparing the ensemble model to all other models combined in 2023/24 (top) and 2024/25 (bottom) winter seasons. **(B)** Distribution of relative AE of the ensemble across all countries for each epidemiological week separately (2023/24 in orange, left half; 2024/25 in purple, right half of the split violin). **(C)** Distribution of relative AE of the ensemble across all epidemiological weeks for each country separately (2023/24 in yellow, top half; 2024/25 in green, bottom half of the split violin).. **(D)** Heatmap of relative AE values of the ensemble by country and epidemiological week, with blue indicating better performance (relative AE greater than 0) and red indicating worse performance (relative AE smaller than 0) compared to the baseline. The top-left half of each cell shows data for the 2023/24 season, while the bottom-right half shows data for 2024/25. Cells with grey stripes indicate missing data. The violin plots in Panels B and C are truncated at the observed minimum and maximum values, with dashed lines inside representing the 1st quartile, median, and 3rd quartile of the distribution. In all panels, relative AE values are averaged over the four-week forecasting horizon.

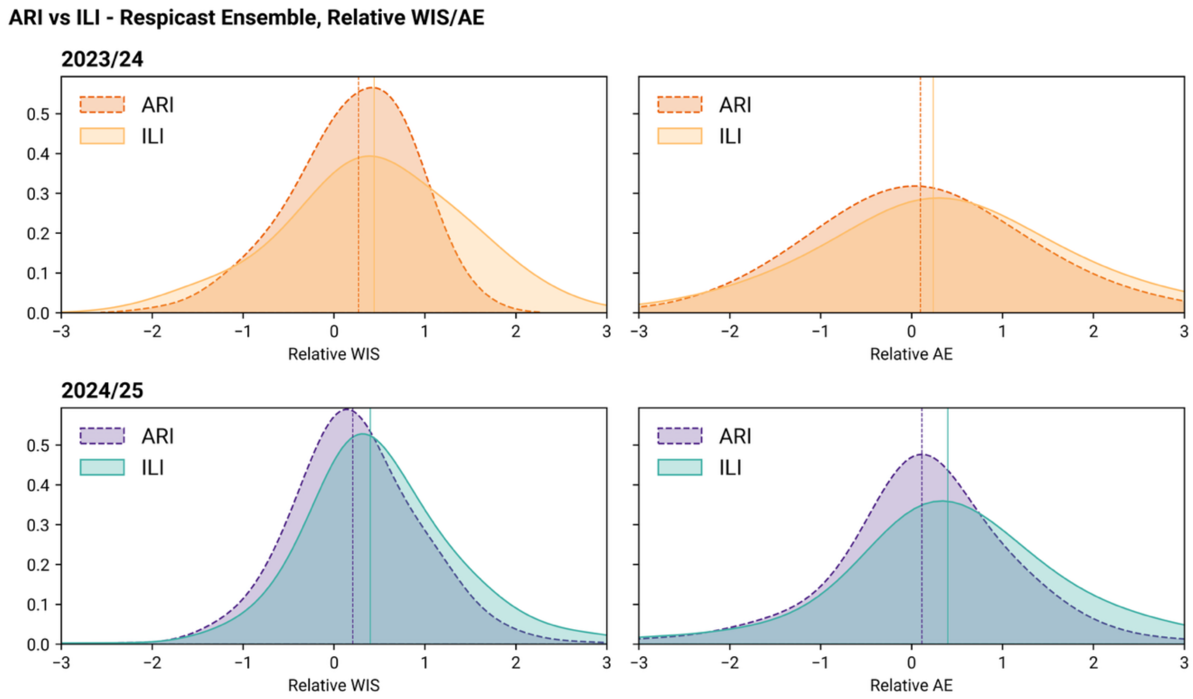


**Figure S14.** Comparison of ensemble forecasts performance for ILI and ARI incidence in 2023/24 (top row) and 2024/25 (bottom row) winter seasons. Left subplots show Relative Weighted Interval Score (WIS) of the ensemble across all countries and forecasting rounds for ILI and ARI incidence forecasting target. Right subplots show Absolute Error (AE) of the median of the ensemble across all countries and forecasting rounds for ILI and ARI incidence forecasting target.

#### S3.2 Respicast ensemble performance compared to individual submitting models

We compare the performance of the RespiCast ensemble to that of individual models in terms of relative WIS and AE for ILI and ARI incidence in each of the two seasons considered. Here, relative performance is computed with respect to the RespiCast ensemble rather than the baseline, meaning that positive values indicate better performance compared to the ensemble, while negative values indicate worse performance. Since different models contributed forecasts for varying numbers of countries and forecasting rounds, we ensure a fair comparison by considering only the rounds common to each model and the ensemble. Additionally, in this comparison we only consider models that contributed to at least 50% of the rounds in each season. This leads to the exclusion of 2 models for ARI incidence in 2023/24, and 3 models for both targets in 2024/25. Table S2 reports the number of forecasting rounds each model participated in for each target and season.

Figure S15 compares the performance of the RespiCast Ensemble to individual models in terms of relative WIS and AE for ILI incidence. The violin plots show the distribution of performance relative to the ensemble based on common rounds. For relative WIS (left) in 2023/24 (top row), no individual model exhibits a positive median, indicating that the ensemble outperforms all individual models for this metric in median terms. When considering relative AE (right), only one model (*EpiEKF*) achieves a positive median relative AE of 0.08, suggesting a modest improvement over the RespiCast ensemble. In 2024/25, we find similar patterns, with only one model showing comparable median performance to that of the ensemble (*FluABCaster*, with a relative WIS and AE of, respectively, 0.01 and 0.005). Figure S16 presents the same analysis for ARI incidence. In this case, no models show a median positive relative WIS or AE in both seasons, indicating that, in median terms, the ensemble outperforms all individual models across different scoring rules and forecasting seasons.

Overall, these results highlight the advantage of using an ensemble over individual models. However, while the ensemble generally outperforms individual models, there are instances where specific models surpass it in certain forecasting rounds, as reflected in the portions of the distribution that extend into the positive domain in Figures S15 and S16.

To better understand the performance of the RespiCast ensemble model compared to individual submitting models, Figures S17 and S18 show the distribution of WIS and AE rank scores for each model and season, for ILI and ARI incidence targets, respectively. For model $m$ and instance $i$ (i.e., forecast round) the rank score is defined as:

$$rank_{m,i}^{score} = 1 - \frac{rank_{m,i} - 1}{N_i - 1} \,,$$

where $N_i$ denotes the total number of submitting models in instance $i$, and $rank_{m,i}$ is the rank of model $m$ for that instance according to the evaluation metric (either WIS or AE). It follows that a rank score of 1 (0) indicates the best (worst) model.

Figure S17 reports the rank score distribution of each model (aggregated across all countries and forecasting rounds) for ILI incidence. In 2023/24, the RespiCast ensemble achieves the highest median WIS rank score, while ranking third for AE (noting that *metaFlu* only submitted forecasts for Italy). In 2024/25, the ensemble is the top-performing model for both WIS and AE median rank scores. Beyond median values, the ensemble's rank score distributions are markedly narrower, indicating more consistent performance across rounds and countries. Although not always the best in every country and round combination, the ensemble most frequently ranks among the top models and least often among the bottom ones (its first quartile of rank scores is always the highest). This consistency is particularly desirable in epidemic forecasting for public health, where robustness and reliability are valued as much as absolute performance.

Figure S18 presents analogous results for ARI incidence. In 2023/24, the ensemble ranks second in terms of both median WIS and AE rank scores, again showing a much narrower distribution than any individual model. In 2024/25, it ranks first for median WIS and second for median AE, with the highest first quartile and smallest IQR among all models.

Finally, Figure S19 summarises the distribution of model performance across seasons by jointly considering how often each model falls in the bottom 30% and top 30% of the WIS ranking. We use the median values along both axes (computed across all targets and seasons) to define four performance quadrants. In the top-left quadrant, we find models that consistently achieve a low proportion of bottom-ranked forecasts while frequently appearing among the top performers. The ensemble falls in this quadrant for all seasons and targets. In all cases, it is also the model that least frequently (almost never) appears in the bottom 30%.

In the 2024/25 season, it is additionally the model that most frequently appears in the top 30%. In 2023/24 for ILI incidence, the ensemble is surpassed by a single model with higher frequency of top-30% placements, although this model submitted forecasts only for Italy, making direct comparison less straightforward. In the 2023/24 ARI case, only one model shows a higher frequency of top-30% appearances, but at the cost of a substantially higher frequency in the bottom 30%. Most remaining models tend to concentrate around the antidiagonal, indicating that those appearing more frequently in the top 30% also tend to appear less often in the bottom 30%, and vice versa, although some deviations from this pattern are visible. Few models cluster in the bottom-left quadrant (particularly in 2024/25), reflecting stable middle performance (i.e., models that rarely feature among the best performers but also avoid the worst outcomes). Other models fall in the top-right quadrant, especially in 2023/24. We label these models as volatile performers, as they frequently appear in both the top and bottom 30%. Finally, models consistently underperforming relative to the rest appear in the bottom-right quadrant.

Figure S20 shows analogous results using AE-based rankings. The findings are broadly similar, although the ensemble's performance is weaker than under WIS. It remains within the *High Performers* quadrant in all cases and continues to be the model least frequently appearing in the bottom 30%. However, its frequency of top-30% appearances is reduced across all panels and is surpassed by a few individual models in some instances, though these models always exhibit a higher proportion of bottom-30% placements.

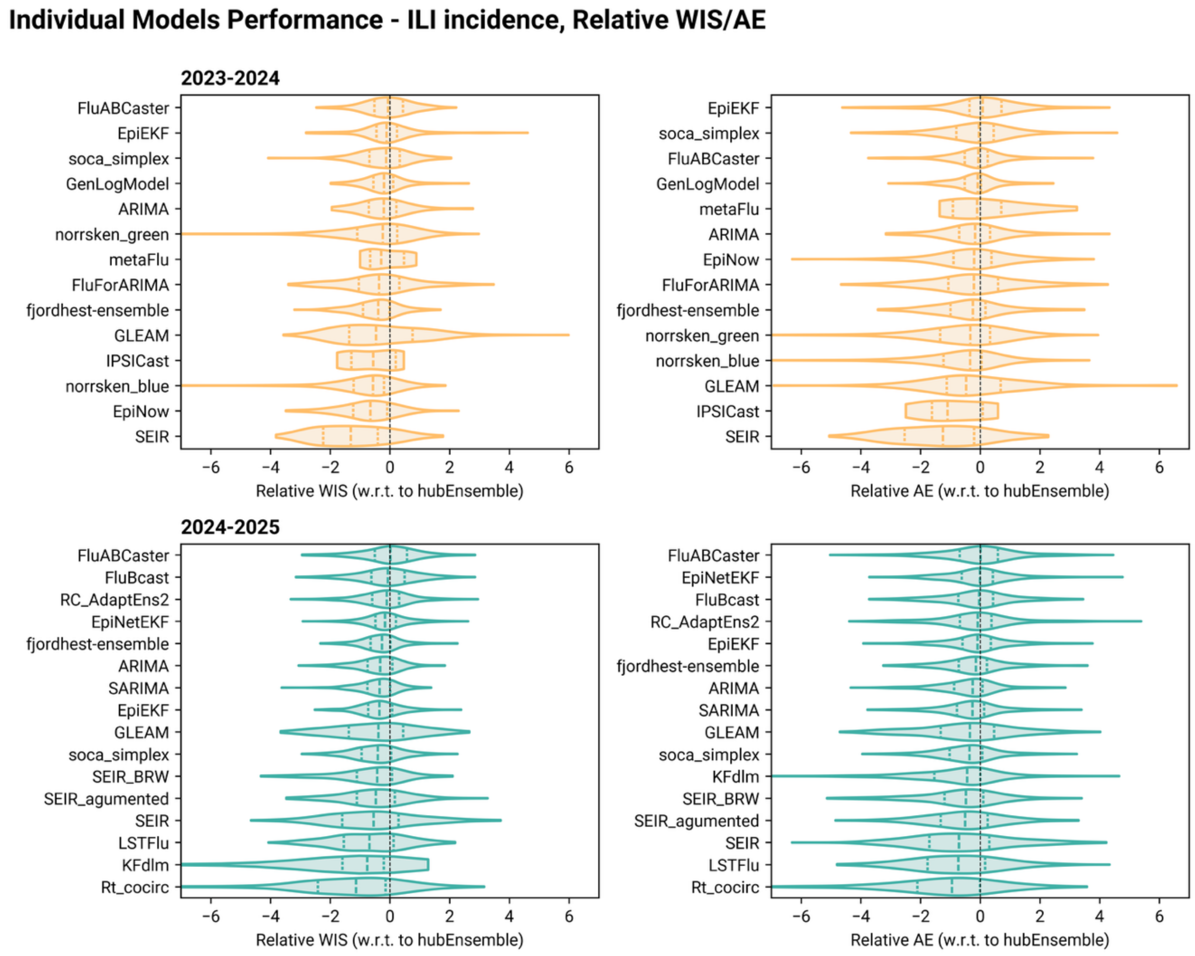


**Figure S15.** Performance comparison of individual models for ILI incidence in terms of relative Weighted Interval Score (WIS) (left) and relative Absolute Error (AE) (right) during the 2023/24 (top row) and 2024/25 (bottom row) seasons. Each violin plot represents the distribution of relative scores for a given model with respect to the ensemble benchmark (hubEnsemble). Negative values indicate worse performance relative to the ensemble, while positive values indicate better performance. Dashed lines inside the violins represent the 1st quartile, median, and 3rd quartile of the distribution. Only countries and forecasting rounds that are common to both each model and the ensemble have been considered. Additionally, only models that submitted forecasts in at least half of the rounds in each season are displayed.

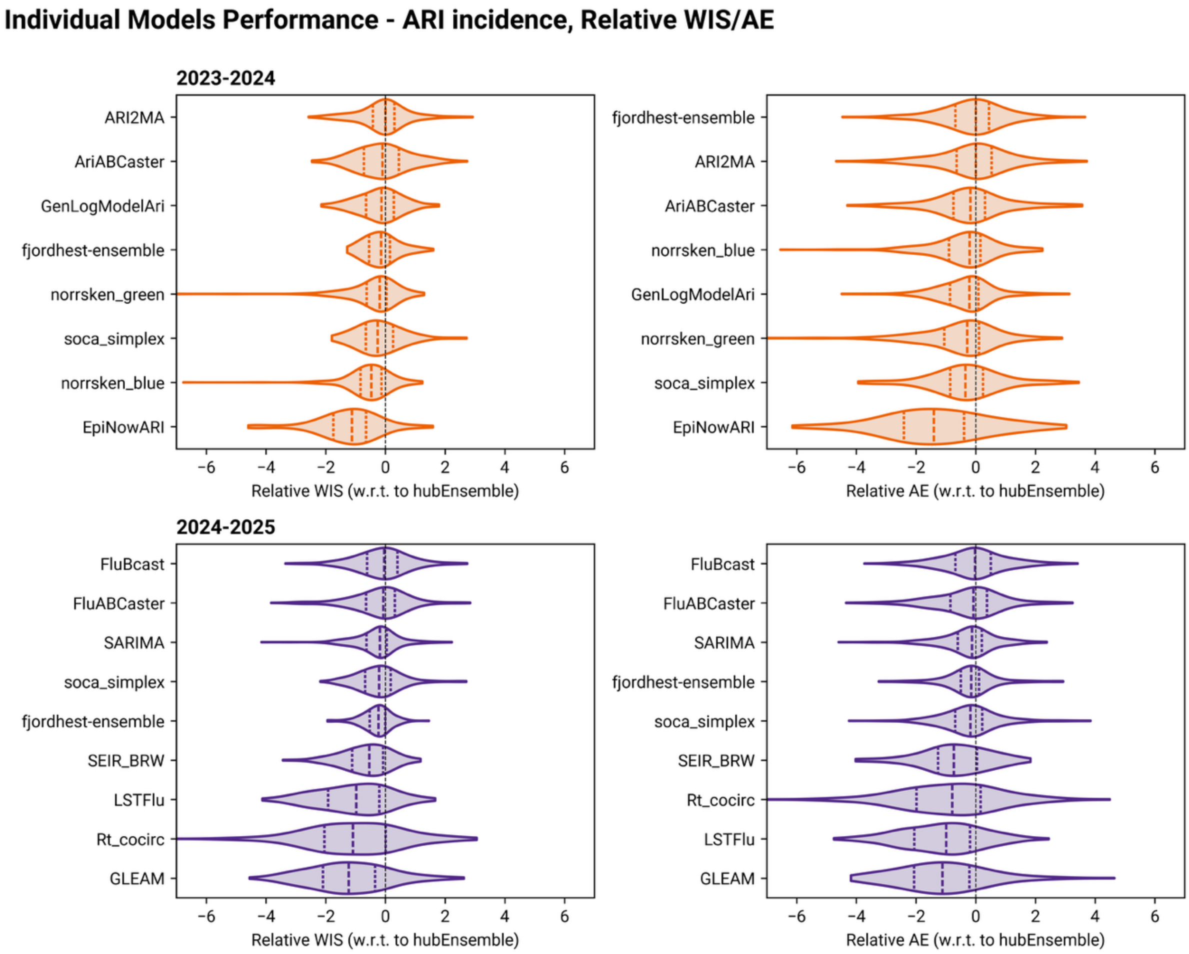


**Figure S16.** Performance comparison of individual models for ARI incidence in terms of relative Weighted Interval Score (WIS) (left) and relative Absolute Error (AE) (right) during the 2023/24 (top row) and 2024/25 (bottom row) seasons. Each violin plot represents the distribution of relative scores for a given model with respect to the ensemble benchmark (hubEnsemble). Negative values indicate worse performance relative to the ensemble, while positive values indicate better performance. Dashed lines inside the violins represent the 1st quartile, median, and 3rd quartile of the distribution. Only countries and forecasting rounds that are common to both each model and the ensemble have been considered. Additionally, only models that submitted forecasts in at least half of the rounds in each season are displayed.

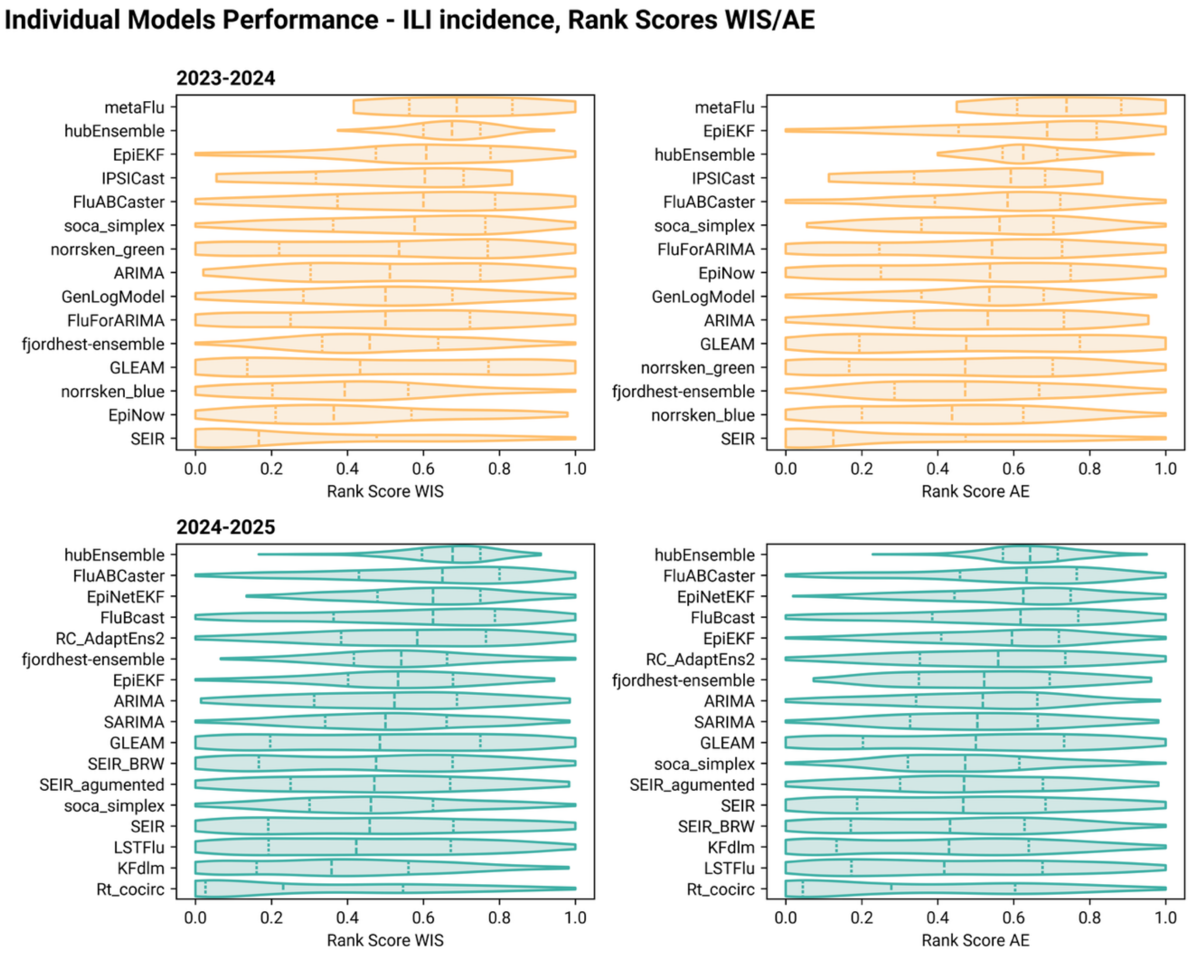


Figure S17. Performance comparison of individual models for ILI incidence in terms of WIS (left) and AE (right) rank score during the 2023/24 (top row) and 2024/25 (bottom row) seasons. Each violin plot represents the distribution of rank scores for a given model, scoring rule, and season. Higher (lower) values of rank score indicate better (worse) performance with respect to other models. Only models that submitted forecasts in at least half of the rounds in each season are displayed.

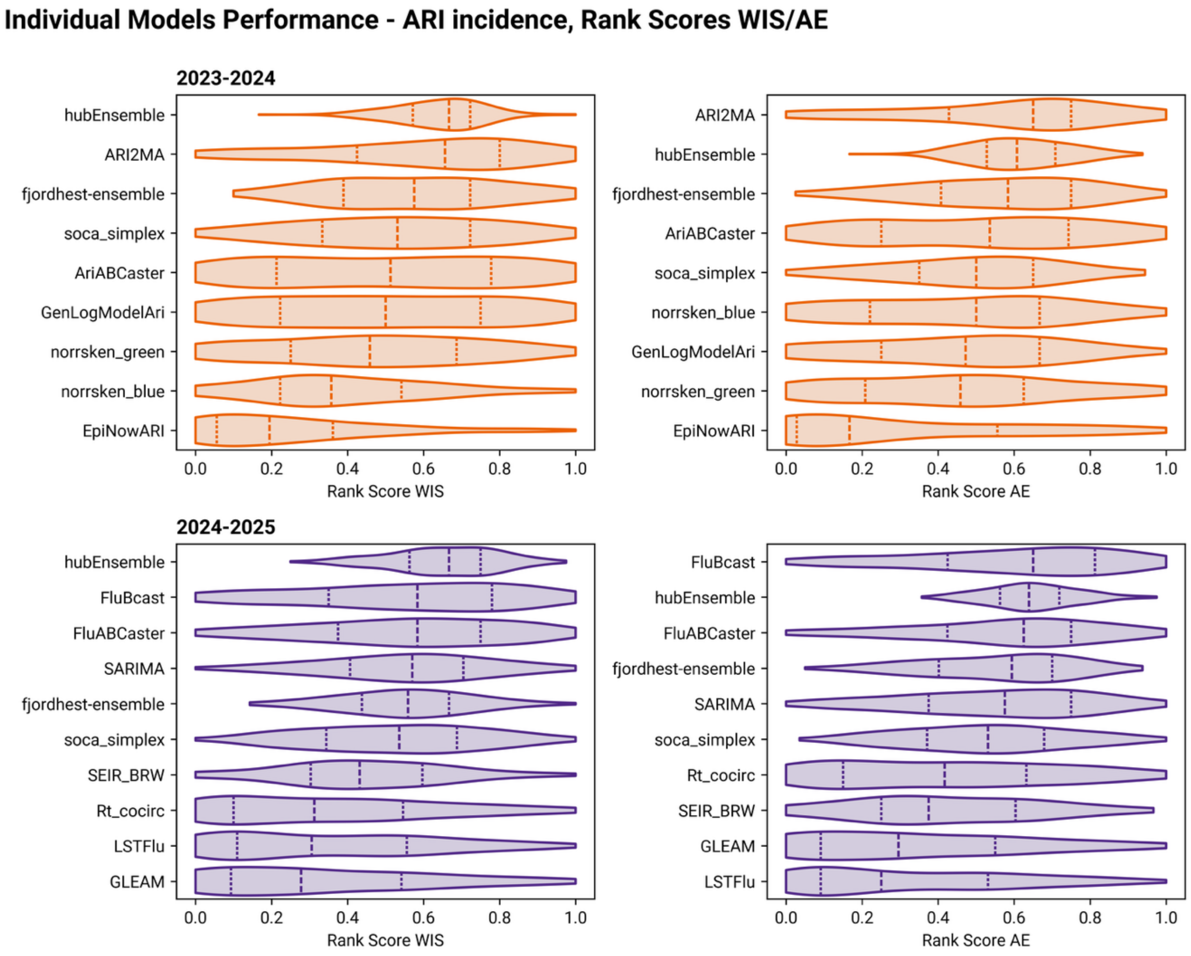


Figure S18. Performance comparison of individual models for ARI incidence in terms of WIS (left) and AE (right) rank score during the 2023/24 (top row) and 2024/25 (bottom row) seasons. Each violin plot represents the distribution of rank scores for a given model, scoring rule, and season. Higher (lower) values of rank score indicate better (worse) performance with respect to other models. Only models that submitted forecasts in at least half of the rounds in each season are displayed.

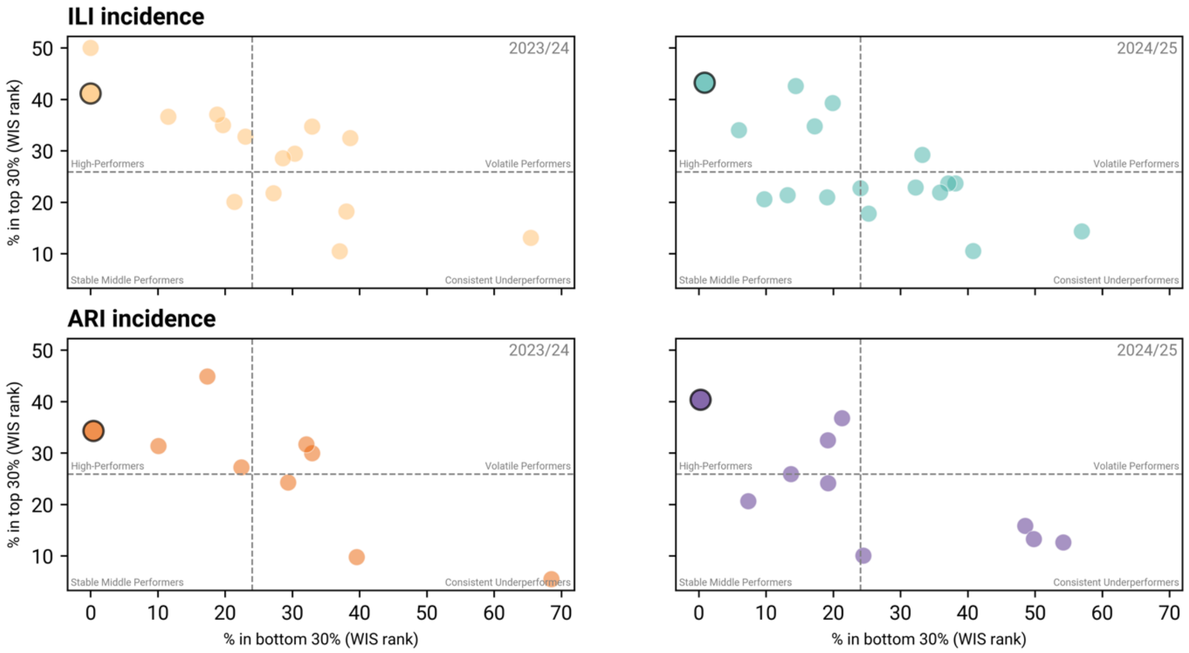


Figure S19. Performance distribution of individual models and the ensemble for ILI and ARI incidence forecasts across seasons 2023/24 and 2024/25 (WIS). Each point represents a model, with the ensemble highlighted. The x-axis shows the percentage of forecasting rounds in which a model ranked in the bottom 30%, and the y-axis shows the percentage of rounds in which it ranked in the top 30%. Quadrants are defined using the median values of both axes across all models, seasons, and targets, yielding four performance categories: High performers (low bottom-30% frequency, high top-30% frequency), Volatile performers (high frequency in both extremes), Stable middle performers (low frequency in both extremes), and Consistent underperformers (high bottom-30% frequency, low top-30% frequency).

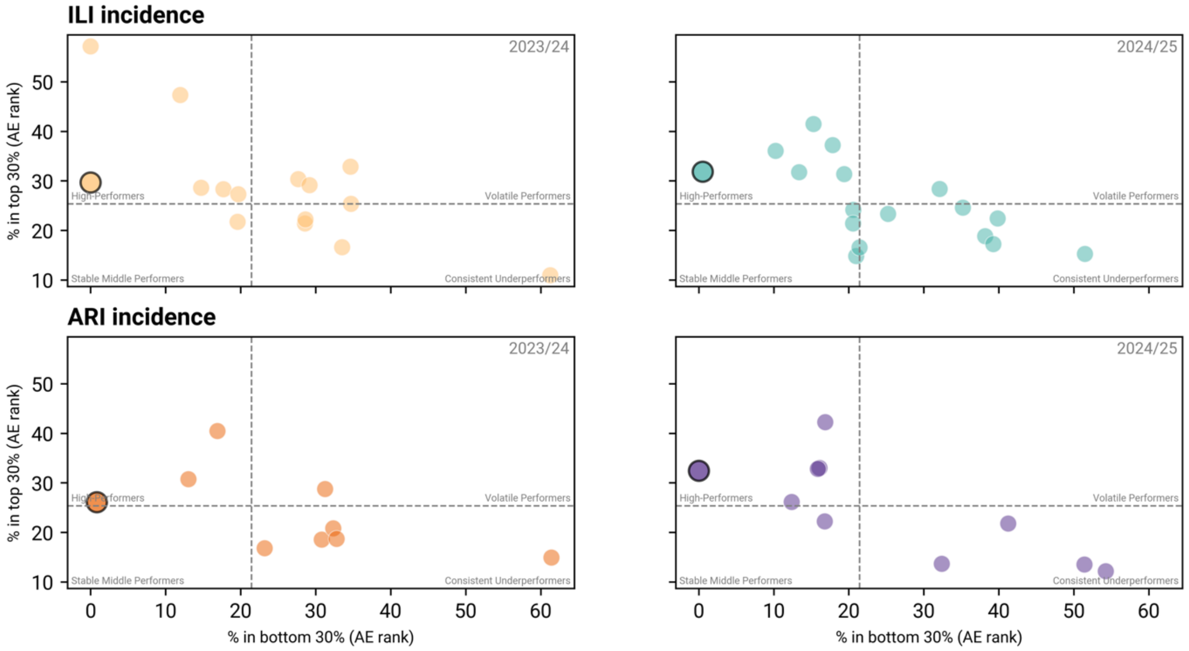


Figure S20. Performance distribution of individual models and the ensemble for ILI and ARI incidence forecasts across seasons 2023/24 and 2024/25 (AE). Each point represents a model, with the ensemble highlighted. The x-axis shows the percentage of forecasting rounds in which a model ranked in the bottom 30%, and the y-axis shows the percentage of rounds in which it ranked in the top 30%. Quadrants are defined using the median values of both axes across all models, seasons, and targets, yielding four performance categories: High performers (low bottom-30% frequency, high top-30% frequency), Volatile performers (high frequency in both extremes), Stable middle performers (low frequency in both extremes), and Consistent underperformers (high bottom-30% frequency, low top-30% frequency).

#### S3.3 Prediction coverage

Figure S21 illustrates the 90% coverage deviations (defined as the difference between nominal and prediction coverage) for the ensemble model across locations for both ILI and ARI incidence forecasting target in the two seasons. Deviations below 0 indicate under-coverage, reflecting overconfidence in the forecasts, while deviations near 0 suggest well-calibrated predictions. The ILI results (top row) show considerable heterogeneity, with deviations ranging from approximately -0.4 to 0.1 in both seasons, indicating substantial under-coverage in many locations, particularly in Croatia, Romania, and Slovenia in 2023/24 and Belgium, Luxembourg, and Norway in 2023/24. Conversely, locations like Italy and Poland in 2023/24 or Austria and Iceland in 2024/25 demonstrate better-calibrated forecasts, with deviations much closer to 0. For ARI forecasts (bottom row), the coverage deviations follow similar trends, with overall undercoverage patterns in both seasons and strong heterogeneity across locations.

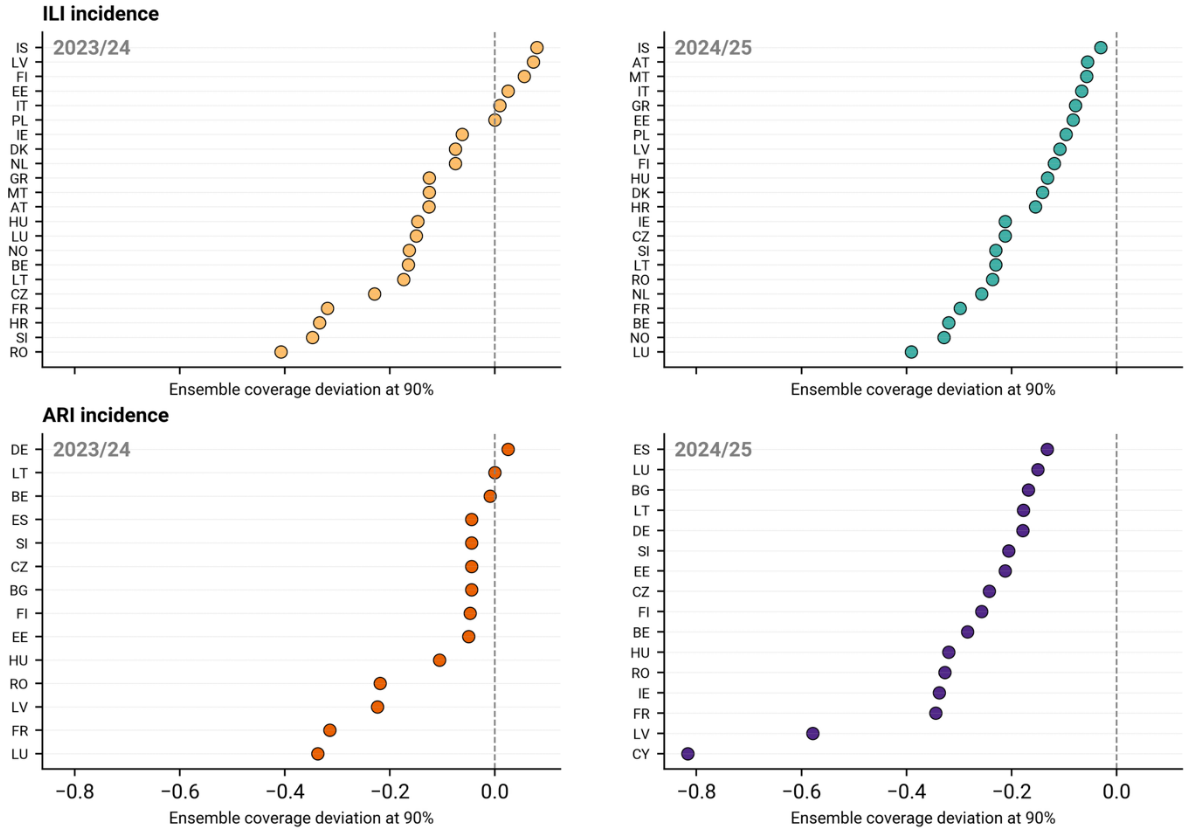


**Figure S21.** Ensemble 90% coverage deviations for ILI (top row) and ARI (bottom row) incidence in 2023/24 (left) and 2024/25 (right) seasons across locations. Each point represents the deviation between the observed 90% prediction interval coverage and the nominal 90% coverage for a specific location, target, and season. Negative deviations indicate under-coverage (overconfidence), positive deviations indicate over-coverage (underconfidence), while values close to 0 indicate well-calibrated forecasts.

#### S3.4 Sensitivity analysis for relative performance computation

In the main text, we computed relative WIS on a per-round basis by taking the ratio of baseline to model WIS for each forecasting round separately, then aggregating these ratios across rounds for visualization and analysis. To assess the robustness of our main findings to alternative relative performance definition, we conducted a sensitivity analysis using the pairwise comparison approach proposed in Ref. [1], which addresses potential concerns about the order of operations in computing relative performance metrics.

The procedure consists of the following steps, performed independently for each combination of target (ILI or ARI incidence), season (2023/24 or 2024/25), country, and forecasting horizon (1 to 4 weeks ahead):

1. For each pair of models $(m, m')$ we identify the set of forecasting rounds for which both models submitted forecasts. We then compute the mean absolute WIS for each model over these common rounds and compute their ratio:
$$\theta_{m,m'} = \frac{WIS_{avg}^{m}}{WIS_{avg}^{m'}}$$
2. For each model $m$, we compute the geometric mean of its pairwise scores across all other models:
$$\theta_m = \left(\prod_{m'=1}^{M} \theta_{m,m'}\right)^{1/M},$$
where M is the total number of models that submitted forecasts for the given target, season, country, and horizon combination.

3. We compute the relative WIS for each model $m$ by dividing its $\theta_m$by that of the baseline:
$$\theta_m^* = \frac{\theta_m}{\theta_B}$$

The obtained quantity $\theta_m^*$ is the relative WIS of model $m$, adjusted for the difficulty of the forecasts it produced and rescaled such that the baseline model has $\theta_B^* = 1$. It follows that, $\theta_m^* < 1$indicates improved performance of model $m$ with respect to the baseline. We note that, in this sensitivity analysis, WIS values are computed on log-transformed incidence data prior to aggregation [2]. This transformation reduces the disproportionate influence of high-incidence periods on aggregate performance metrics, facilitating more direct comparison with the relative performance metrics presented in the main text. Analogous steps can be applied to compute relative AE.

Figure S22 presents the distribution of relative WIS and AE computed using this alternative methodology, separately for ILI (left panels) and ARI incidence (right panels). Each boxplot shows the distribution of relative scores across countries for a given season and forecasting horizon (the *Combined* category represents relative scores obtained by pooling all horizons together). Values smaller than 1.0 (indicated by the horizontal dashed line) indicate that the ensemble outperforms the baseline model.

The sensitivity analysis confirms the principal findings presented in the main text. Indeed, the ensemble consistently achieves relative WIS smaller than 1.0, across most horizons, countries, and seasons, confirming that it outperforms the baseline regardless of the aggregation method used.

For ILI incidence, in both seasons, the median relative WIS ranges from approximately 0.6 to 0.8 for different horizons, with substantial variability across countries (as indicated by the boxplot ranges). During the 2023/24 season, when considering the results of all horizons combined, only three countries feature an ensemble relative WIS greater than 1. In 2024/25 the ensemble achieves a relative WIS smaller than 1 in all countries. For ARI incidence, relative WIS distributions are generally closer to 1, indicating more modest performance gains over the baseline compared to ILI incidence, as discussed also in the main text. For this target, the ensemble outperforms the baseline in all but five countries in each season.

For specific targets and seasons, relative WIS values tend to be lower at longer forecasting horizons, suggesting a greater performance advantage for the ensemble over the baseline as the forecast horizon increases. However, this trend is neither strong nor consistent across all settings, indicating that no single horizon consistently dominates when aggregating results across countries and epidemic phases.

Similar patterns emerge when evaluating relative performance using the absolute error of the median forecast, even though the performance advantage is more modest compared to WIS-based metrics, especially for ARI incidence, consistent with the findings presented in the main text and above.

Finally, to assess consistency between the two methodologies, we compared the ensemble relative WIS/AE of different countries using both the per-round approach (main text) and the pairwise approach (this sensitivity analysis). We find statistically significant correlations across all target-season combinations using both Kendall and Spearman correlation coefficients (Table S3), indicating strong agreement between methods regarding which countries exhibit better or worse ensemble performance relative to the baseline. The negative sign of the correlation coefficients is expected given our definition of relative WIS of the main text (where higher values indicate better performance) in contrast with the definition used in the sensitivity analysis (where lower values indicate better performance). Nonetheless, we stress that these negative correlations indicate positive agreement between methods.

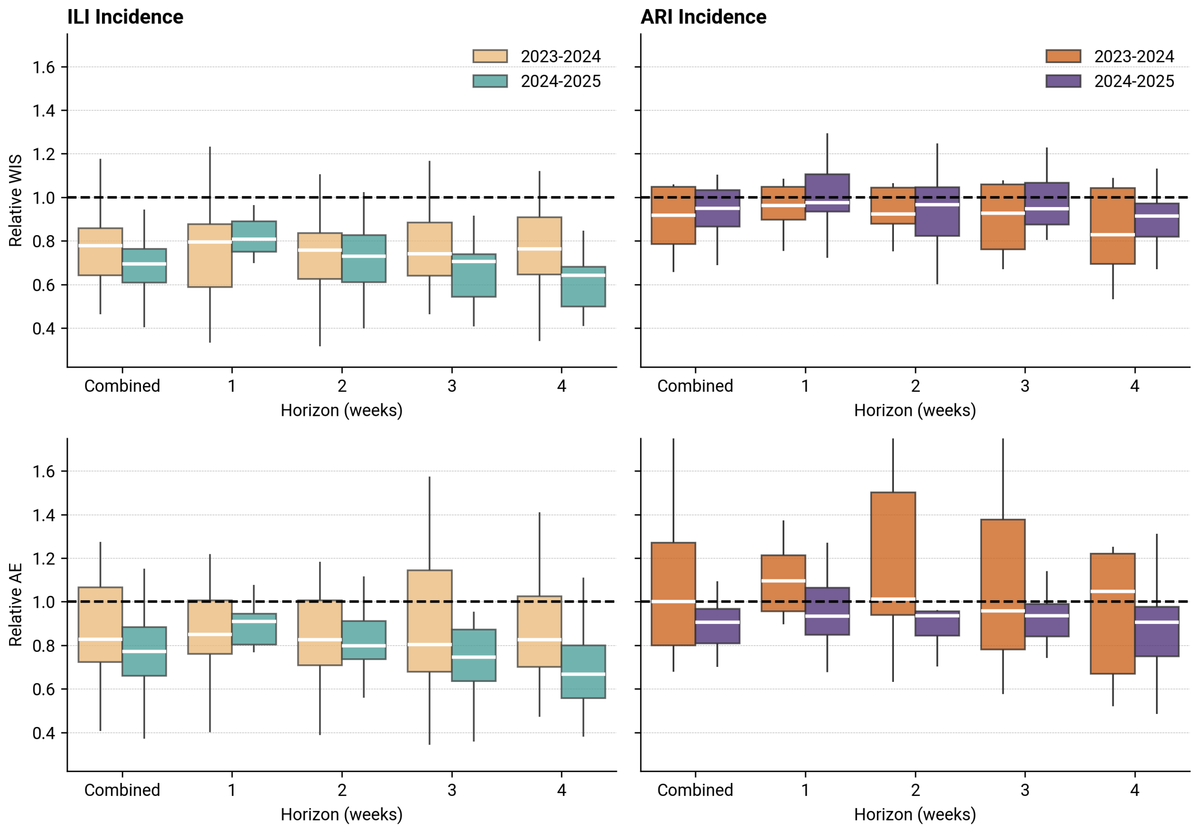


**Figure S22.** Boxplots of ensemble relative WIS (top) and AE (bottom) computed as in Ref. [1] for ILI (left) and ARI (right) incidence at the country level. In each panel, relative scores are shown separately for each season and forecasting horizon. Values below 1 indicate improved performance with respect to the baseline.

| | | ILI Incidence | | ARI Incidence | |
|---|---|---|---|---|---|
| | | Kendall $\tau$ | Spearman $\rho$ | Kendall $\tau$ | Spearman $\rho$ |
| 2023/24 | Rel. WIS | -0.69 (***) | -0.84 (***) | -0.52 (**) | -0.70 (**) |
| | Rel. AE | -0.63 (***) | -0.80 (***) | -0.69 (***) | -0.86 (***) |
| 2024/25 | Rel. WIS | -0.49 (**) | -0.65 (**) | -0.67 (***) | -0.84 (***) |
| | Rel. AE | -0.55 (***) | -0.75 (***) | -0.38 (*) | -0.44 (*) |

Table S3. Correlation between relative WIS and AE as computed in the main text (i.e., median per-round relative WIS) and as computed in the sensitivity analysis (i.e., pairwise comparison from Ref. [1]). Correlation is computed considering both the Kendall and Spearman coefficients for each season and target. Significance is displayed as follows: *** indicates $p_{val} < 0.001$, ** $p_{val} < 0.01$, * indicates $p_{val} < 0.05$, blank otherwise. Negative correlations are expected due to the definitions of relative WIS but still indicates positive agreement.
